# Concurrent RB1 loss and *BRCA*-deficiency predicts enhanced immunological response and long-term survival in tubo-ovarian high-grade serous carcinoma

**DOI:** 10.1101/2023.11.09.23298321

**Authors:** Flurina A. M. Saner, Kazuaki Takahashi, Timothy Budden, Ahwan Pandey, Dinuka Ariyaratne, Tibor A. Zwimpfer, Nicola S. Meagher, Sian Fereday, Laura Twomey, Kathleen I. Pishas, Therese Hoang, Adelyn Bolithon, Nadia Traficante, Kathryn Alsop, Elizabeth L. Christie, Eun-Young Kang, Gregg S. Nelson, Prafull Ghatage, Cheng-Han Lee, Marjorie J. Riggan, Jennifer Alsop, Matthias W. Beckmann, Jessica Boros, Alison H. Brand, Angela Brooks-Wilson, Michael E. Carney, Penny Coulson, Madeleine Courtney-Brooks, Kara L. Cushing-Haugen, Cezary Cybulski, Mona A. El-Bahrawy, Esther Elishaev, Ramona Erber, Simon A. Gayther, Aleksandra Gentry-Maharaj, C. Blake Gilks, Paul R. Harnett, Holly R. Harris, Arndt Hartmann, Alexander Hein, Joy Hendley, AOCS Group, Brenda Y. Hernandez, Anna Jakubowska, Mercedes Jimenez-Linan, Michael E. Jones, Scott H. Kaufmann, Catherine J. Kennedy, Tomasz Kluz, Jennifer M. Koziak, Björg Kristjansdottir, Nhu D. Le, Marcin Lener, Jenny Lester, Jan Lubiński, Constantina Mateoiu, Sandra Orsulic, Matthias Ruebner, Minouk J. Schoemaker, Mitul Shah, Raghwa Sharma, Mark E. Sherman, Yurii B. Shvetsov, Naveena Singh, T. Rinda Soong, Helen Steed, Paniti Sukumvanich, Aline Talhouk, Sarah E. Taylor, Robert A. Vierkant, Chen Wang, Martin Widschwendter, Lynne R. Wilkens, Stacey J. Winham, Michael S. Anglesio, Andrew Berchuck, James D. Brenton, Ian Campbell, Linda S. Cook, Jennifer A. Doherty, Peter A. Fasching, Renée T. Fortner, Marc T. Goodman, Jacek Gronwald, David G. Huntsman, Beth Y. Karlan, Linda E. Kelemen, Usha Menon, Francesmary Modugno, Paul D.P. Pharoah, Joellen M. Schildkraut, Karin Sundfeldt, Anthony J. Swerdlow, Ellen L. Goode, Anna DeFazio, Martin Köbel, Susan J. Ramus, David D. L. Bowtell, Dale W. Garsed

**Affiliations:** Peter MacCallum Cancer Centre, Melbourne, Victoria, Australia; Department of Obstetrics and Gynecology, Bern University Hospital and University of Bern, Bern, Switzerland; Department of Obstetrics and Gynecology, The Jikei University School of Medicine, Tokyo, Japan; School of Clinical Medicine, UNSW Medicine and Health, University of NSW Sydney, Sydney, New South Wales, Australia; Skin Cancer and Ageing Lab, Cancer Research United Kingdom Manchester Institute, The University of Manchester, Manchester, UK; The Daffodil Centre, The University of Sydney, a joint venture with Cancer Council NSW, Sydney, New South Wales, Australia; Sir Peter MacCallum Department of Oncology, The University of Melbourne, Parkville, Victoria, Australia; Adult Cancer Program, Lowy Cancer Research Centre, University of NSW Sydney, Sydney, New South Wales, Australia; Department of Pathology and Laboratory Medicine, University of Calgary, Foothills Medical Center, Calgary, AB, Canada; Department of Oncology, Division of Gynecologic Oncology, Cumming School of Medicine, University of Calgary, Calgary, AB, Canada; Department of Laboratory Medicine and Pathology, University of Alberta, Edmonton, Alberta, Canada; Department of Obstetrics and Gynecology, Division of Gynecologic Oncology, Duke University Medical Center, Durham, NC, USA; Centre for Cancer Genetic Epidemiology, Department of Oncology, University of Cambridge, Cambridge, UK; Department of Gynecology and Obstetrics, Comprehensive Cancer Center Erlangen-EMN, Friedrich-Alexander University Erlangen-Nuremberg, University Hospital Erlangen, Erlangen, Germany; Centre for Cancer Research, The Westmead Institute for Medical Research, Sydney, New South Wales, Australia; Department of Gynaecological Oncology, Westmead Hospital, Sydney, New South Wales, Australia; The University of Sydney, Sydney, New South Wales, Australia; Canada’s Michael Smith Genome Sciences Centre, BC Cancer, Vancouver, BC, Canada; Department of Obstetrics and Gynecology, John A. Burns School of Medicine, University of Hawaii, Honolulu, HI, USA; Division of Genetics and Epidemiology, The Institute of Cancer Research, London, UK; Department of Obstetrics, Gynecology and Reproductive Sciences, University of Pittsburgh School of Medicine, Pittsburgh, PA, USA; Program in Epidemiology, Division of Public Health Sciences, Fred Hutchinson Cancer Center, Seattle, WA, USA; Department of Genetics and Pathology, International Hereditary Cancer Center, Pomeranian Medical University, Szczecin, Poland; Department of Metabolism, Digestion and Reproduction, Imperial College London, Hammersmith Hospital, London, UK; Department of Pathology, University of Pittsburgh School of Medicine, Pittsburgh, PA, USA; Institute of Pathology, Comprehensive Cancer Center Erlangen-EMN, Friedrich-Alexander University Erlangen-Nuremberg, University Hospital Erlangen, Erlangen, Germany; Center for Bioinformatics and Functional Genomics and the Cedars Sinai Genomics Core, Cedars-Sinai Medical Center, Los Angeles, CA, USA; MRC Clinical Trials Unit, Institute of Clinical Trials and Methodology, University College London, London, UK; Department of Women’s Cancer, Elizabeth Garrett Anderson Institute for Women’s Health, University College London, London, UK; Department of Pathology and Laboratory Medicine, University of British Columbia, Vancouver, BC, Canada; Crown Princess Mary Cancer Centre, Westmead Hospital, Sydney, New South Wales, Australia; Department of Epidemiology, University of Washington, Seattle, WA, USA; QIMR Berghofer Medical Research Institute, Brisbane, Queensland, Australia; University of Hawaii Cancer Center, Honolulu, HI, USA; Independent Laboratory of Molecular Biology and Genetic Diagnostics, Pomeranian Medical University, Szczecin, Poland; Department of Histopathology, Addenbrooke’s Hospital, Cambridge, UK; Division of Oncology Research, Department of Oncology, Mayo Clinic, Rochester, MN, USA; Department of Gynecology and Obstetrics, Gynecology Oncology and Obstetrics, Institute of Medical Sciences, Medical College of Rzeszow University, Rzeszów, Poland; Alberta Health Services-Cancer Care, Calgary, AB, Canada; Department of Obstetrics and Gynecology, Institute of Clinical Sciences, Sahlgrenska Center for Cancer Research, University of Gothenburg, Gothenburg, Sweden; Cancer Control Research, BC Cancer Agency, Vancouver, BC, Canada; International Hereditary Cancer Center, Department of Genetics and Pathology, Pomeranian Medical University in Szczecin, Szczecin, Poland; David Geffen School of Medicine, Department of Obstetrics and Gynecology, University of California at Los Angeles, Los Angeles, CA, USA; Department of Pathology, University of Gothenburg, Gothenburg, Sweden; Tissue Pathology and Diagnostic Oncology, Westmead Hospital, Sydney, New South Wales, Australia; Department of Health Sciences Research, Mayo Clinic, Jacksonville, FL, USA; Division of Gynecologic Oncology, Department of Obstetrics and Gynecology, University of Alberta, Edmonton, Alberta, Canada; Section of Gynecologic Oncology Surgery, North Zone, Alberta Health Services, Edmonton, Alberta, Canada; British Columbia’s Gynecological Cancer Research Team (OVCARE), University of British Columbia, BC Cancer, and Vancouver General Hospital, Vancouver, BC, Canada; Department of Obstetrics and Gynecology, University of British Columbia, Vancouver, BC, Canada; Department of Quantitative Health Sciences, Division of Clinical Trials and Biostatistics, Mayo Clinic, Rochester, MN, USA; Department of Quantitative Health Sciences, Division of Computational Biology, Mayo Clinic, Rochester, MN, USA; EUTOPS Institute, University of Innsbruck, Innsbruck, Austria; Cancer Research UK Cambridge Institute, University of Cambridge, Cambridge, UK; Epidemiology, School of Public Health, University of Colorado, Aurora, CO, USA; Community Health Sciences, University of Calgary, Calgary, AB, Canada; Huntsman Cancer Institute, Department of Population Health Sciences, University of Utah, Salt Lake City, UT, USA; Division of Cancer Epidemiology, German Cancer Research Center (DKFZ), Heidelberg, Germany; Department of Research, Cancer Registry of Norway, Oslo, Norway; Cancer Prevention and Control Program, Cedars-Sinai Cancer, Cedars-Sinai Medical Center, Los Angeles, CA, USA; Department of Molecular Oncology, BC Cancer Research Centre, Vancouver, BC, Canada; Division of Acute Disease Epidemiology, South Carolina Department of Health & Environmental Control, Columbia, SC, USA; Department of Epidemiology, University of Pittsburgh School of Public Health, Pittsburgh, PA, USA; Women’s Cancer Research Center, Magee-Womens Research Institute and Hillman Cancer Center, Pittsburgh, PA, USA; Department of Computational Biomedicine, Cedars-Sinai Medical Center, West Hollywood, CA, USA; Centre for Cancer Genetic Epidemiology, Department of Public Health and Primary Care, University of Cambridge, Cambridge, UK; Department of Epidemiology, Rollins School of Public Health, Emory University, Atlanta, GA, USA; Division of Breast Cancer Research, The Institute of Cancer Research, London, UK; Department of Quantitative Health Sciences, Division of Epidemiology, Mayo Clinic, Rochester, MN, USA

## Abstract

**Background:** Somatic loss of the tumour suppressor RB1 is a common event in tubo-ovarian high-grade serous carcinoma (HGSC), which frequently co-occurs with alterations in homologous recombination DNA repair genes including *BRCA1* and *BRCA2* (*BRCA*). We examined whether tumour expression of RB1 was associated with survival across ovarian cancer histotypes (HGSC, endometrioid (ENOC), clear cell (CCOC), mucinous (MOC), low-grade serous carcinoma (LGSC)), and how co-occurrence of germline *BRCA* pathogenic variants and RB1 loss influences long-term survival in a large series of HGSC.

**Patients and methods:** RB1 protein expression patterns were classified by immunohistochemistry in epithelial ovarian carcinomas of 7436 patients from 20 studies participating in the Ovarian Tumor Tissue Analysis consortium and assessed for associations with overall survival (OS), accounting for patient age at diagnosis and FIGO stage. We examined RB1 expression and germline *BRCA* status in a subset of 1134 HGSC, and related genotype to survival, tumour infiltrating CD8+ lymphocyte counts and transcriptomic subtypes. Using CRISPR-Cas9, we deleted *RB1* in HGSC cell lines with and without *BRCA1* mutations to model co-loss with treatment response. We also performed genomic analyses on 126 primary HGSC to explore the molecular characteristics of concurrent homologous recombination deficiency and *RB1* loss.

**Results:** RB1 protein loss was most frequent in HGSC (16.4%) and was highly correlated with *RB1* mRNA expression. RB1 loss was associated with longer OS in HGSC (hazard ratio [HR] 0.74, 95% confidence interval [CI] 0.66-0.83, *P* = 6.8 × 10^-7^), but with poorer prognosis in ENOC (HR 2.17, 95% CI 1.17-4.03, *P* = 0.0140). Germline *BRCA* mutations and RB1 loss co-occurred in HGSC (*P* < 0.0001). Patients with both RB1 loss and germline *BRCA* mutations had a superior OS (HR 0.38, 95% CI 0.25-0.58, *P* = 5.2 x10^-6^) compared to patients with either alteration alone, and their median OS was three times longer than non-carriers whose tumours retained RB1 expression (9.3 years vs. 3.1 years). Enhanced sensitivity to cisplatin (*P* < 0.01) and paclitaxel (*P* < 0.05) was seen in *BRCA1* mutated cell lines with *RB1* knockout. Among 126 patients with whole-genome and transcriptome sequence data, combined *RB1* loss and genomic evidence of homologous recombination deficiency was correlated with transcriptional markers of enhanced interferon response, cell cycle deregulation, and reduced epithelial-mesenchymal transition in primary HGSC. CD8+ lymphocytes were most prevalent in *BRCA*-deficient HGSC with co-loss of *RB1*.

**Conclusions:** Co-occurrence of RB1 loss and *BRCA* mutation was associated with exceptionally long survival in patients with HGSC, potentially due to better treatment response and immune stimulation.

## INTRODUCTION

Despite a high response rate to primary treatment, the progressive development of acquired drug resistance is common in tubo-ovarian high-grade serous carcinoma (HGSC), a histotype that is associated with approximately 70% of ovarian cancer deaths^1^. The frequent acquisition of resistance-conferring alterations in HGSC^2–4^ suggests that the development of drug resistance may be inevitable when curative surgery is not achieved in these patients. Countering that view, however, is the observation that a small subset of patients with HGSC advanced disease experience an exceptional response to treatment, survive well beyond a median of 3.4 years^5^, and in some cases, remain disease free^6,7^. Interest in studying long-term cancer survivors is growing as they may assist the discovery of prognostic biomarkers, novel treatments, and approaches to limit the development of resistance^8^.

Several clinical and molecular factors that influence treatment response and overall survival (OS) in HGSC have been described. Complete surgical debulking is associated with a more favourable outcome compared to patients left with residual disease^9–11^. Molecular subtypes defined by distinct gene expression patterns in primary HGSC are associated with different outcomes^12^, including the poor survival C1/mesenchymal subtype that is more often seen in patients where complete surgical tumour resection cannot be achieved^13–15^. By contrast, the C2/immunoreactive subtype is typified by extensive infiltration of intraepithelial T cells^12^, a feature known to be strongly associated with improved survival^16,17^. Tumours arising in individuals with germline or somatic alterations in *BRCA1* or *BRCA2* genes are typically more responsive to conventional chemotherapy and poly(ADP-ribose) polymerase (PARP) inhibitors, whereas those tumours with intact homologous recombination (HR) DNA repair are more often resistant to treatment^18–20^. Patients with germline *BRCA1* or *BRCA2* pathogenic variants show more favourable survival at five years post-diagnosis compared to non-carriers, with *BRCA2* mutation carriers retaining a long-term (>10 year) survival advantage^21–23^. Although deleterious mutations in *BRCA1*, *BRCA2* and other genes involved in HR DNA repair are associated with a favourable response to treatment, these are not sufficient alone to confer long-term survival and a large proportion of such patients experience a typical disease trajectory. A differential outcome in mutation carriers can in part be ascribed to alternative splicing^24^ or retention of the wild-type *BRCA* allele in tumours^25^, both of which appear to limit the effectiveness of chemotherapy.

We previously characterised a small series of HGSC exceptional survivors and found that co-occurring loss of function alterations in both *BRCA* and *RB1* were associated with unusually favourable survival^7,26^. Disruption of the RB pathway is found in many cancer types but with variable impacts on patient outcome. For example, co-loss of *RB1* and *BRCA* is associated with shorter survival in breast and prostate cancer, possibly due to lineage switching and resistance to hormonal therapy^27–29^. A transcriptomic signature of RB1 loss was recently described to be associated with poor outcomes across cancer types^30^. We have previously found that chromosomal breakage is the most common mechanism of *RB1* inactivation in HGSC^3^, accounting for approximately 80% of all *RB1* alterations. In addition to its crucial role in cell cycle regulation, RB1 is involved in non-canonical functions in a context- and tissue-dependent manner^31–33^, including HR mediated DNA repair. Loss of RB1 expression in HGSC has been associated with a survival benefit^34^, including in the context of abnormal block-like p16 staining^35^.

Factors underlying the association of RB1 loss with improved outcome in HGSC are unknown. Here, we contrast the pattern and clinical consequences of RB1 loss in HGSC with other epithelial ovarian cancer subtypes, investigate the relevance of co-occurring *BRCA1* or *BRCA2* mutations and RB1 loss in HGSC patients, and explore the functional effects of combined *BRCA* and *RB1* impairment in HGSC cell lines.

## PATIENTS AND METHODS

### Patient cohorts

The study population consisted of 7436 patients diagnosed with invasive epithelial ovarian, peritoneal or fallopian tube cancer from 20 studies or biobanks participating in the Ovarian Tumor Tissue Analysis (OTTA) consortium^36^ (Supplementary Fig. S1). Written informed consent or IRB approved waiver of consent was obtained at each site for patient recruitment, sample collection, and study protocols (Supplementary Table S1).

Whole-genome sequence and matched transcriptome sequence data of primary HGSC tumours were available from 126 patients from the Multidisciplinary Ovarian Cancer Outcomes Group (MOCOG) study^26^ (Supplementary Fig. S1). This cohort consisted of 34 short-term survivors (OS <2 years), 32 moderate-term survivors (OS ≥2 and <10 years) and 60 long-term survivors (OS ≥10 years) with advanced stage (IIIC/IV) disease, enrolled in the Australian Ovarian Cancer Study (AOCS), the Gynaecological Oncology Biobank at Westmead Hospital (Sydney) or the Mayo Clinic Study.

### Molecular analyses

RB1 protein expression was determined by immunohistochemistry (IHC) staining and scoring of tissue microarrays (TMAs) from formalin-fixed paraffin-embedded (FFPE) tumour samples, using our previously described protocol^7^ (RB1 antibody clone 13A10, Leica Biosystems; Supplementary Material). Subsets of HGSC patients had additional molecular or immune data available (Supplementary Fig. S1), including tumour p53 protein expression status previously classified^37^ as normal (wild-type) or abnormal (overexpression, complete absence, and cytoplasmic), germline *BRCA1* and *BRCA2* pathogenic variant status obtained from OTTA, *RB1* mRNA tumour expression obtained using NanoString (ref^34^ and unpublished data), transcriptional subtypes of tumours using NanoString^38^ and CD8+ tumour infiltrating lymphocyte (TIL) density was previously classified^39^ based on the number of CD8+ TILs per high-powered field: negative (no TILs), low (<3 TILs), moderate (3-19 TILs) or high (≥20 TILs).

The MOCOG whole-genome and transcriptome sequencing dataset of 126 short-, moderate- and long-term survivors was uniformly processed as previously described^26^, and included detailed characterisation of each tumour sample for inactivating alterations in *RB1* and HR pathway genes, including germline and/or somatic mutations in *BRCA1*, *BRCA2*, *BRIP1*, *PALB2*, *RAD51C* and *RAD51D*, or promoter methylation of *BRCA1* and *RAD51C*. Homologous recombination deficiency (HRD) status was assessed using the CHORD (Classifier of Homologous Recombination Deficiency) method^40^, which uses specific base substitution, indel and structural rearrangement signatures detected in tumour genomes to generate *BRCA1*-type and *BRCA2*-type HRD scores. Primary tumours were classified as either *BRCA1*-HRD & *RB1* altered; *BRCA1*-HRD & *RB1* wild-type; *BRCA2*-HRD & *RB1* altered; *BRCA2*-HRD & *RB1* wild-type; homologous recombination proficient (HRP) & *RB1* altered, or HRP & *RB1* wild-type. For details on differential gene expression analyses, see Supplementary Material.

### Cell culture

The AOCS patient-derived cell lines (AOCS1, AOCS3, AOCS7.2 AOCS9, AOCS11.2, AOCS14, AOCS16, AOCS22, AOCS30) were established from ascites drained from patients with HGSC, as previously described^4^. All AOCS cell lines were authenticated against matched patient germline DNA using short tandem repeat markers (STR, GenePrint10 System, Promega). Commercial cell lines OAW28 and CAOV3, categorised as likely HGSC^41^, were purchased from the American Type Culture Collection (ATCC), and JHOS2 and OVCAR4 were obtained from the National Cancer Institute Repository. Commercial lines were authenticated by comparing STR profiles (GenePrint10 System, Promega) to those published by online repositories (Cancer Cell Line Encyclopaedia, The Cancer Genome Atlas) before use in experiments. Cell lines were confirmed to be free of *Mycoplasma* by PCR at each revival and after finishing experiments. For details on cell growth conditions, CRISPR-mediated gene knockout, and molecular and functional cell line characterisation, see Supplementary Material.

### Statistical analyses

Cox proportional hazards models were used to estimate hazard ratios (HRs) with 95% confidence intervals (CIs) using the ‘coxph’ function of the R package *survival* (v3.2-7). Final models were fitted using Cox regression adjusted for age at diagnosis and FIGO stage. A spline function was used for age at diagnosis with degree of freedom (df) 5 to account for the non-linear effect of the continuous variable. Regression models were fitted separately by histotype. The HGSC regression models were also stratified by site of participant recruitment, and sites with fewer than 10 events within the study period were excluded. The ENOC regression model was not stratified by site due to the limited number of overall patients per site. The OTTA survival dataset was right censored at 10 years from diagnosis to reduce the number of non-ovarian cancer related deaths. In the final Cox regression model, there was evidence for deviation from the proportional hazard assumption, but the degree of deviation was not substantial when considered alongside the large sample size and Schoenfeld residuals. The Kaplan–Meier method was used to estimate and plot progression-free and overall survival probabilities, and the log-rank (Mantel–Cox) test used to compare the survival duration between subgroups. In the Kaplan-Meier curves, the number of patients at risk on the date of diagnosis (time = 0) may be fewer than subsequent time intervals, owing to left truncation of follow-up resulting from delayed study enrolment at some OTTA sites. Differences in proportions of categorical features were assessed by either the chi-square or Fisher’s exact test as indicated. Differences in continuous variables were assessed by either a Wilcoxon Rank Sum Test or a Kruskal-Wallis test. All *in vitro* assays were performed across at least three independent experiments, and data are expressed as mean ± standard error of the mean (SEM) as indicated, from a minimum of three independent measurements. All statistical tests were two-sided and considered significant when *P* < 0.05. Statistical analyses were performed using either Prism (v9.3.1) or R (v3.6.3).

## RESULTS

### Loss of RB1 expression is most frequent in HGSC

RB1 protein expression was assessed by IHC in tumour samples from 7436 ovarian cancer patients using TMAs from 20 centres participating in the OTTA consortium (Supplementary Tables S1 and S2). RB1 tumour expression was classified as either retained or lost in 6564 samples, with 872 samples excluded that had either subclonal loss (*n* = 66), cytoplasmic (*n* = 17), or uninterpretable results (*n* = 789) due to either sample drop out or the absence of an internal positive control (Fig. 1A, Supplementary Material).

**Figure 1.**
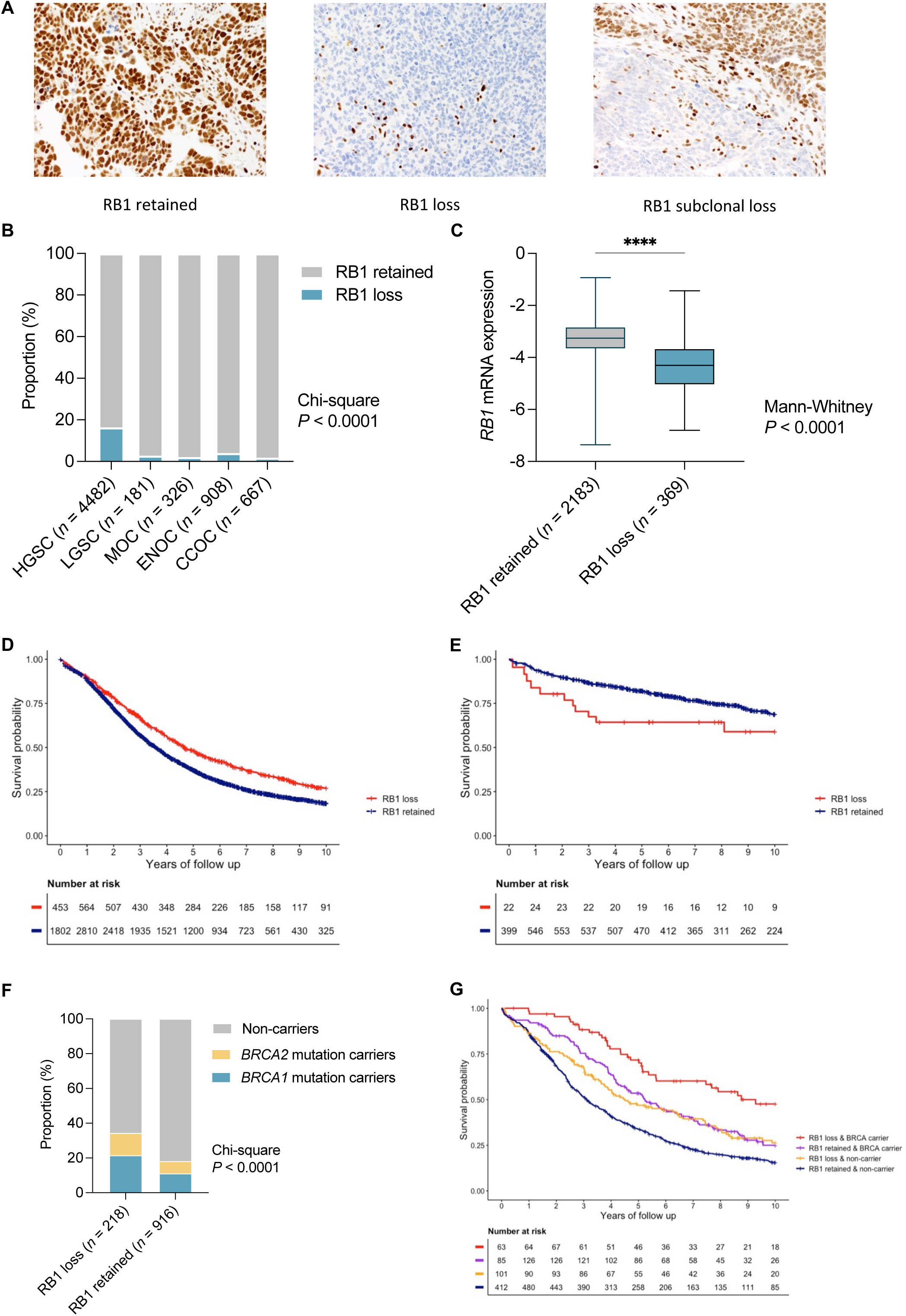
Expression of RB1 and survival associations across ovarian cancer histotypes. (A) Representative images of immunohistochemical detection of RB1 expression in ovarian carcinoma tissues, showing examples of the three most common expression patterns: retained, lost and subclonal loss. (B) Proportion of patients with loss or retention of RB1 protein expression in tumour samples by ovarian cancer histotypes. Chi-square *P* value reported for difference in proportions across all histotypes. HGSC, tubo-ovarian high-grade serous carcinoma; LGSC, low-grade serous carcinoma; MOC, mucinous ovarian cancer; ENOC, endometrioid ovarian cancer; CCOC, clear cell ovarian cancer. (C) Boxplots show *RB1* mRNA expression (NanoString) by RB1 protein expression status; lines indicate median and whiskers show range (Mann-Whitney test *P* value reported). Kaplan-Meier analysis of overall survival in patients diagnosed with HGSC (D) and ENOC (E) stratified by tumour RB1 expression. (F) Loss of RB1 tumour expression is more common in germline *BRCA1* and *BRCA2* mutation carriers than retained RB1 expression. Chi-square *P* value is reported. (G) Kaplan-Meier estimates of overall survival in HGSC patients by combined germline *BRCA* and tumour RB1 expression status.

RB1 loss was most frequent in HGSC (16.4%), followed by endometrioid ovarian cancer (ENOC; 4.1%, Chi-square *P* < 0.0001, Fig. 1B). Loss of RB1 expression was less frequent in all other histotypes (1.8% to 2.8%). *RB1* mRNA expression was also assessed by NanoString in a subset of HGSC tumours (*n* = 2552) and was significantly associated with RB1 protein expression (Fig. 1C, *P* < 0.0001).

### RB1 loss is associated with longer survival in HGSC

Loss of RB1 protein expression was associated with longer OS in patients with HGSC (HR 0.74, 95% CI 0.66-0.83, *P* = 6.8×10^-7^; Table 1) following multivariate analysis adjusting for stage and age at diagnosis and stratified by study. Patients with HGSC were comparable in terms of stage regardless of RB1 loss or retained expression (*P* = 0.9246), however those with RB1 loss had a younger age at diagnosis (median 59 years versus 61 years, *P* = 0.0003; Supplementary Table S3). Median OS was 4.7 years for patients with RB1 loss compared to 3.6 years for those with retained RB1 expression (Fig. 1D).

**Table 1.**
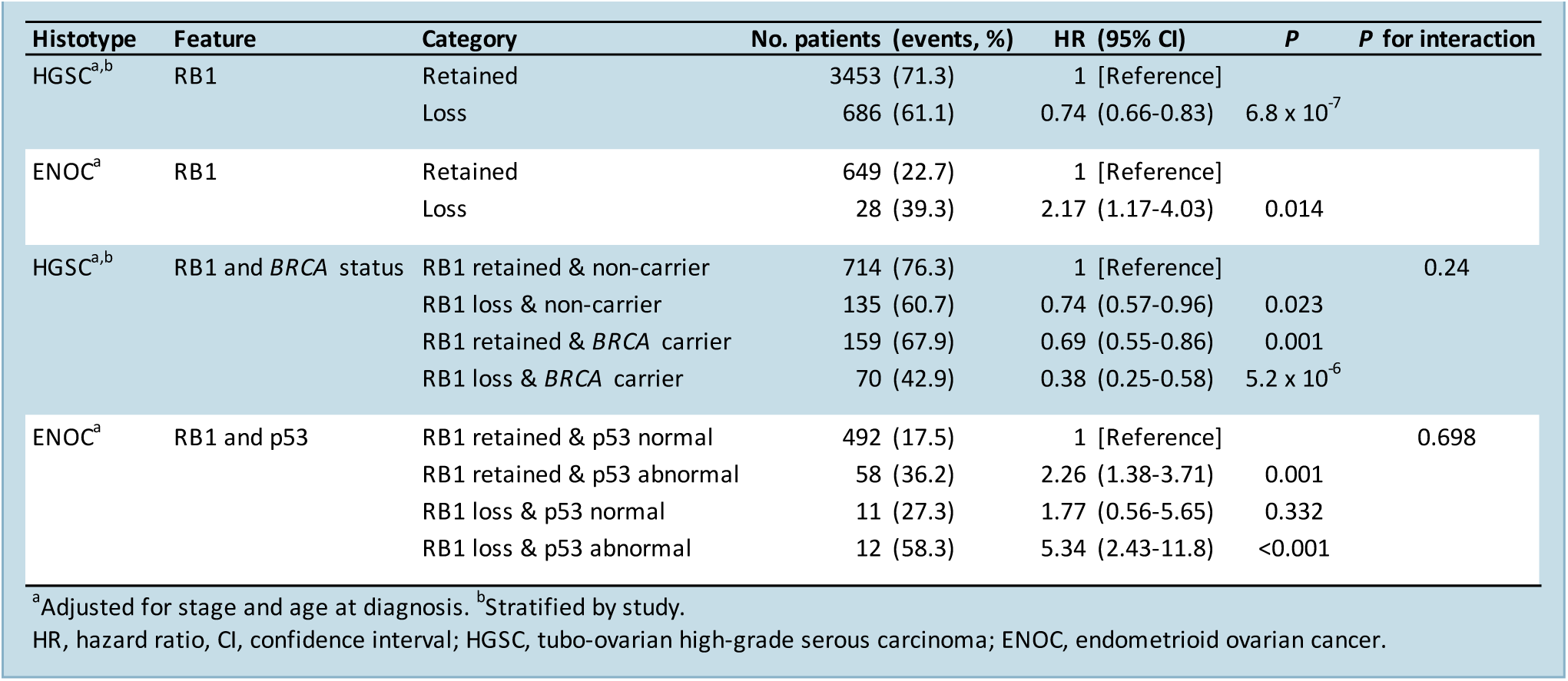
Multivariate analysis of molecular alterations and overall survival in patients with HGSC and ENOC.

In contrast to HGSC, loss of RB1 expression in tumours from patients with ENOC was associated with advanced stage (*P* = 0.0003) and poorer survival (HR 2.17, 95% CI 1.17-4.03, *P* = 0.0140; Table 1, Fig. 1E, Supplementary Table S4). RB1 loss and abnormal p53 protein expression, which is highly predictive of *TP53* mutation^42^, were strongly correlated (chi-square *P* < 0.0001; Supplementary Fig. 2A). While *TP53* mutation is known to be associated with inferior survival in patients with ENOC^37,43^, we note that combined RB1 loss and abnormal p53 expression were associated with the shortest patient survival (median OS 3.0 years; Supplementary Fig. 2B), suggesting that loss of RB1 and *TP53* mutation have a compounding negative impact on survival in patients with ENOC.

### Combined RB1 loss and germline BRCA mutation is associated with exceptionally good survival

We previously observed that co-occurrence of somatic RB1 protein loss and *BRCA1* or *BRCA2* alteration (somatic or germline) was associated with longer progression-free survival (PFS) and OS in HGSC^7^. Here, germline *BRCA1* and *BRCA2* status was available for 1134 HGSC patients for which we had RB1 IHC data (Supplementary Fig. S1). Consistent with having a younger age of diagnosis, patients with RB1 loss were more likely to have concurrent germline *BRCA1* or *BRCA2* mutations than those with retained RB1 expression (Fig. 1F, Chi-square *P* < 0.0001). Patients with both RB1 loss and a germline *BRCA* mutation had a 62% reduced risk of death compared with non-carriers with retained RB1 (HR 0.38, 95% CI 0.25-0.58, *P* = 5.2−10^-6^; Table 1). The median OS of *BRCA* germline carriers with RB1 loss was three times longer than non-carriers with RB1 retained tumours (median OS 9.3 years vs. 3.1 years, respectively), while median OS was 5.2 years for *BRCA* carriers with retained RB1 expression and 4.5 years for non-carriers with RB1 loss (Fig. 1G; Supplementary Table S5).

### Enhanced response to chemotherapy in cells with impaired BRCA and RB1 function

To investigate whether co-occurrence of *RB1* and *BRCA* alterations enhances sensitivity to standard-of-care ovarian cancer drugs, nine patient-derived HGSC cell lines with confirmed pathogenic *TP53* mutation and known *RB1* and *BRCA* status were treated with cisplatin, paclitaxel and olaparib (Supplementary Fig. S3A-C). AOCS14, the only cell line with a germline *BRCA1* mutation and concomitant loss of RB1 expression, showed the best response to cisplatin and olaparib, and was the second most sensitive cell line to paclitaxel. In contrast AOCS11.2, a line with *BRCA1* promoter methylation and loss of RB1 expression, was relatively resistant to paclitaxel and olaparib. Among cell lines with intact RB1 protein expression and *BRCA* wildtype background, AOCS3 was resistant to cisplatin, paclitaxel and olaparib.

Except for the chemo-naïve cell lines AOCS30 and AOCS14, all other lines were derived from patients previously treated with chemotherapy. Since the evaluation of HGSC cell lines with existing *RB1* mutations may have been confounded by their prior, differential exposure to chemotherapy we therefore characterised responses in isogenically matched lines deleted of *RB1* and/or *BRCA1.* We first inactivated *RB1* in two *BRCA1*-mutant (AOCS7.2, AOCS16) and one wild-type line (AOCS1) using CRISPR-Cas9 (Fig. 2A, Supplementary Fig. S4A). *RB1* knockout clones of the *BRCA1*-mutant cell line AOCS7.2 had enhanced sensitivity to cisplatin and paclitaxel compared to *RB1* wild-type clones, which was observed both in short-term drug assays (72 hours, Fig. 2B) and longer-term clonogenic survival assays (12 days, Fig. 2C). In this cell line, sensitivity to paclitaxel and olaparib was increased after *RB1* knockout (paclitaxel IC50 92.0 nM versus 11.8 nM, *P* < 0.0001; olaparib IC50 6.1 versus 1.1 nM, *P* < 0.0001). Further, significantly fewer colonies grew in this *BRCA1*-mutant cell line after *RB1* knockout upon treatment with cisplatin (*P* = 0.01), paclitaxel (*P* = 0.02) or a combination of both drugs (*P* = 0.067) in a clonogenic survival assay (*n* = 3). This effect was not apparent in the *BRCA*-wild-type line (AOCS1) or the other *BRCA1*-mutant line (AOCS16). Western blot and IHC analysis (Supplementary Fig. S4A) found that AOCS16 lacked expression of p16, which may functionally disrupt the RB1 pathway irrespective of an *RB1* knockout^44^.

**Figure 2.**
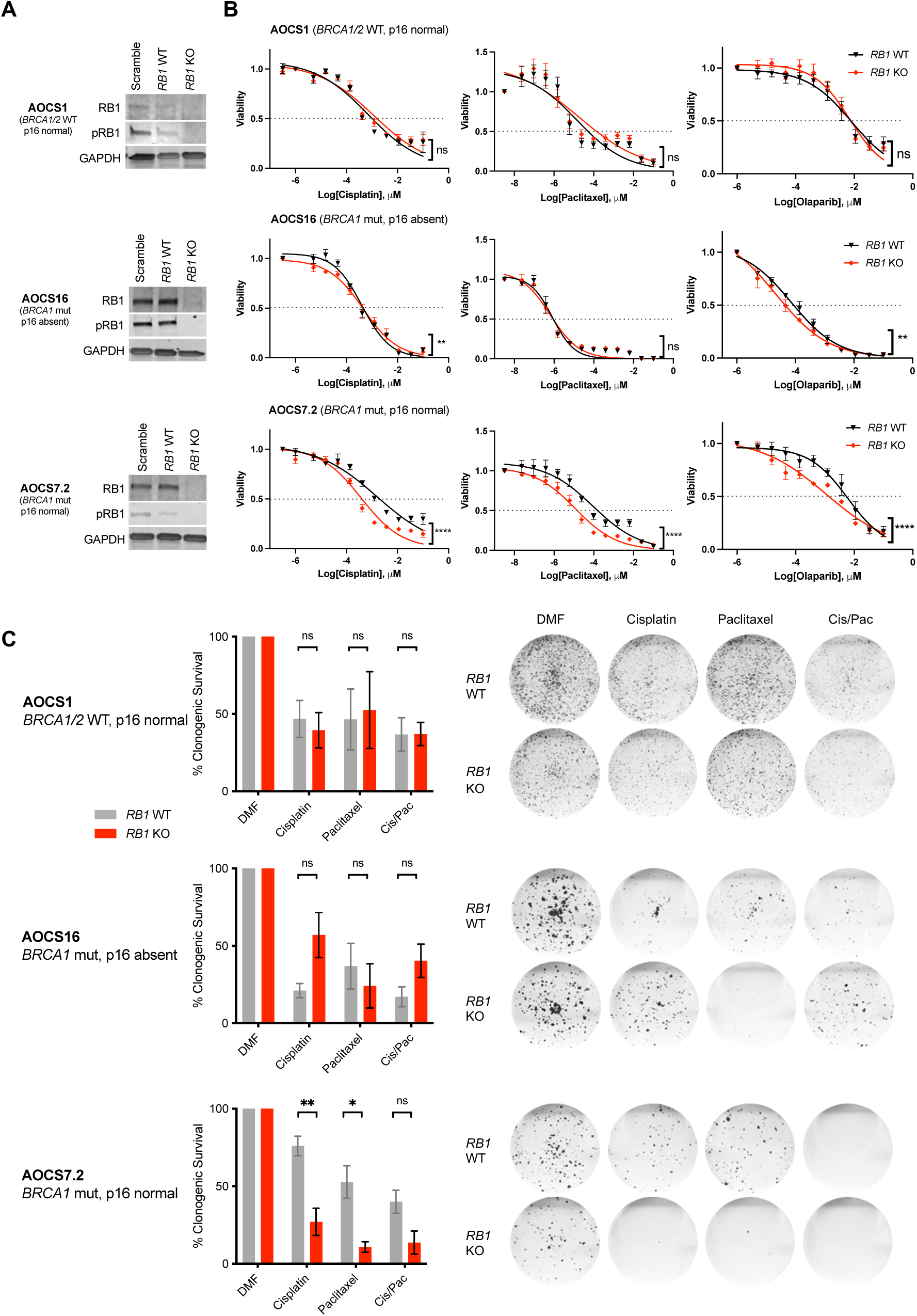
Sensitivity to therapeutic agents in *BRCA1*-mutant cell lines with *RB1* knockout. (A) *RB1* was knocked out using CRISPR/Cas9 in 3 patient-derived Australian Ovarian Cancer Study (AOCS) HGSC cell lines with either wild-type or mutant *BRCA1* background. Representative Western Blots show protein levels of RB1 and phosphorylated RB1 (pRB1) compared to GAPDH loading control in single cell cloned, homozygous *RB1* wildtype (WT) and knockout (KO) colonies in comparison to heterogeneous populations with a scramble single guide RNA (sgRNA). Independent blots were used for RB1 and pRB1. (B) Cell viability was compared between *RB1* WT and KO clones following treatment with cisplatin (72 hours), paclitaxel (72 hours) or olaparib (120 hours). Nonlinear regression drug curves are shown; *P* values of a curve fit, extra sum-of squares F test (ns, not significant; ** *P* < 0.01; **** *P* < 0.0001; *n* = 3). Error bars indicate ± SEM; for some values error bars are shorter than the symbols and thus are not visible. (C) Proportion of surviving colonies following 16 days of treatment with cisplatin, paclitaxel or a combination of both (with half of the IC50 determined per drug and cell line respectively) relative to DMF vehicle control (*n* = 3 replicates). Data are presented as mean ± SEM. Mean values were compared by student’s t-test (ns, not significant; \**P* < 0.05; \*\**P* < 0.01). Representative scans of the fixed cell colonies stained with crystal violet are shown for each condition.

Given that RB1 plays a central role in the negative control of the cell cycle^44,45^, we tested whether the enhanced chemosensitivity of *RB1* knockout AOCS 7.2 cells was associated with increased cell division. Live cell imaging showed similar growth rates of *RB1* wildtype and knockout clones of all three isogenically matched HGSC cell lines (Supplementary Fig. S4B). In both *BRCA* wild-type and *BRCA1* mutant cell lines, *RB1* knockout did not alter cell cycle distribution at baseline or after 24 hours of cisplatin treatment (Supplementary Fig. S4C). Paclitaxel treatment resulted in a larger proportion of cells with a tetraploid DNA content in *RB1* knockout cells compared to *RB1* wild-type cells, indicating arrest in the G2 or M phase of the cell cycle. This effect was observed in all cell lines independent of *BRCA* or p16 status, however the arrest was more profound in the AOCS7.2 cell line (AOCS1, G2/M difference 8.59% ± 4.73%, *P* = 0.144; AOCS16, G2/M difference 8.13% ± 4.45%, *P* = 0.142; AOCS7.2: G2/M difference 14.49% ± 3.99%, *P* = 0.022; Supplementary Fig. S4C).

We extended our analysis of isogenically matched pairs by inactivating *BRCA1* and/or *RB1* in the chemo-naïve cell line AOCS30. While we were readily able to establish *RB1* knockout lines, all *BRCA1* targeted clones were hemizygous for *BRCA1* deletion and retained *BRCA1* expression (Supplementary Table S6), suggesting that engineered homozygous loss of *BRCA1* was cell lethal, even in a tumour type where *BRCA1* loss is frequently observed^46^.

### Genomic and transcriptional landscape of HGSC with combined inactivation of BRCA and RB1

To further understand how RB1 loss may impact the biology of HGSC with co-loss of *BRCA1* or *BRCA2*, we explored matched whole-genome and transcriptome data of primary HGSC tumours in the MOCOG cohort^26^ of 126 short-(OS <2 years), moderate-(OS ≥2 to <10 years) and long-term (OS ≥10 years) survivor patients (Supplementary Fig. S1). Each tumour genome was classified according to their HRD and *RB1* status, resulting in 6 groups: *BRCA1*-HRD & *RB1* altered (*n* = 13); *BRCA1*-HRD & *RB1* wild-type (*n* = 36); *BRCA2*-HRD & *RB1* altered (*n* = 8); *BRCA2*-HRD & *RB1* wild-type (*n* = 20); HRP & *RB1* altered (*n* = 4), or HRP & *RB1* wild-type (*n* = 45; Fig. 3A).

**Figure 3.**
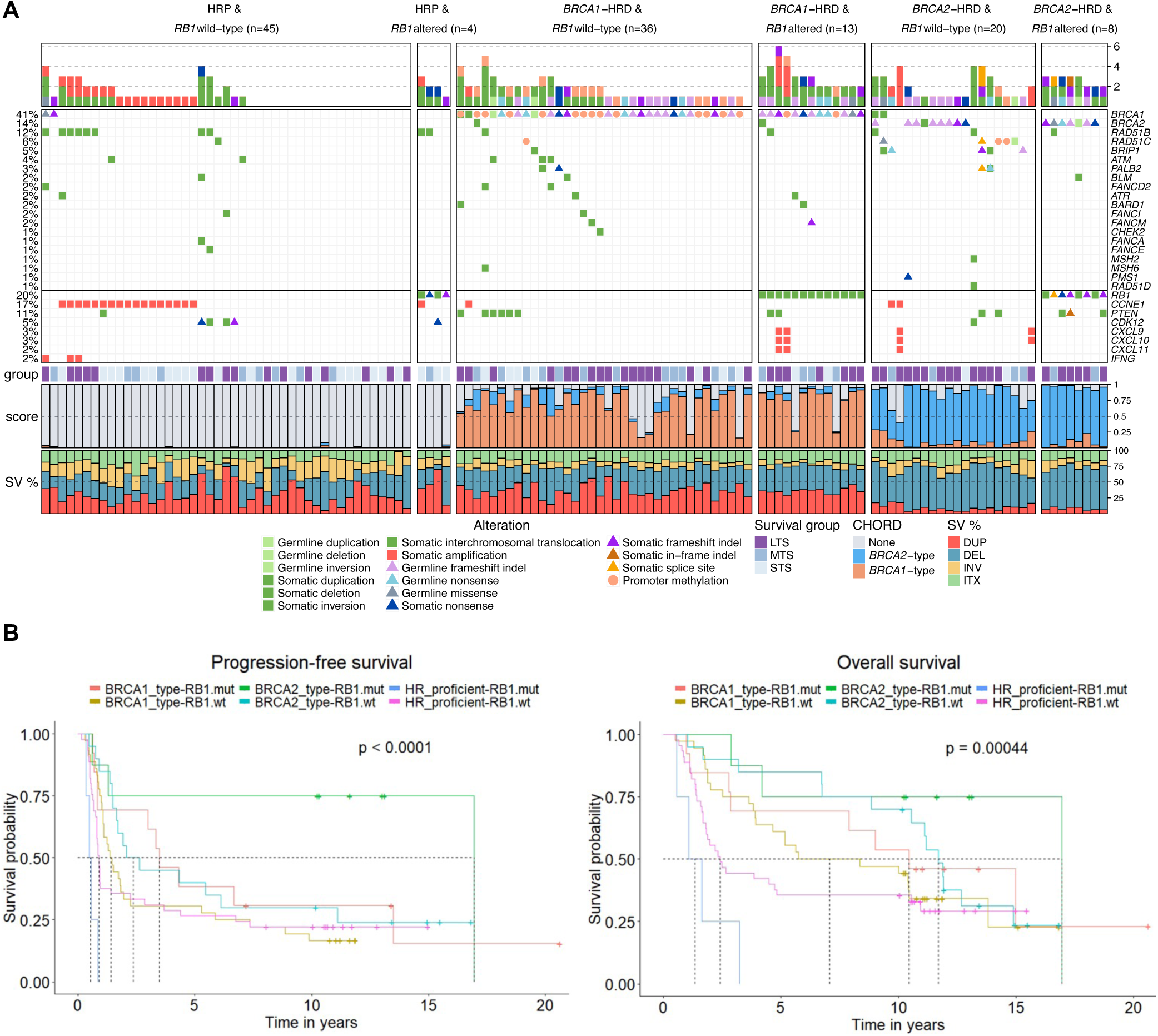
Genomic landscape of high-grade serous ovarian tumours with co-occurring *BRCA* and *RB1* alterations. (A) Pathogenic germline and somatic alterations in homologous recombination (HR) and DNA repair genes detected by whole-genome sequencing and DNA methylation analysis of 126 primary HGSC samples^26^ are shown, as well as alterations in immune genes and *CCNE1*. Samples are grouped by HRD and *RB1* status (wt, wild-type; mut, mutation). Bars at the top indicate the number of alterations in each listed gene per patient. Patients are annotated with survival group (LTS, long-term survivor, OS >10 years; MTS, mid-term survivor, OS 2-10 years; STS, short-term survivor, OS <2 years), tumour CHORD^40^ scores, and the proportion of structural variant (SV) type (DUP, duplication; DEL, deletion; INV, inversion; ITX, intra-chromosomal translocation). (B) Kaplan-Meier estimates of progression-free and overall survival of patients with according to HR status (*BRCA1*-type HRD, *BRCA2*-type HRD or homologous recombination proficient tumours) and *RB1* status (mut, mutation; wt, wild-type).

The cohort had been selected for a long-term survivor study^26^ and hence was enriched for patients with very long survival. Among *BRCA2*-HRD patients, those with *RB1* alterations had longer OS (median OS 17.0 years) compared with those without *RB1* alterations (median OS 11.7 years, *P* = 0.0004; Fig. 3B). Similarly, *BRCA1*-HRD patients with *RB1* alterations survived longer (median OS 10.4 years) than those with an intact *RB1* gene (median OS 7.1 years). There were few HRP tumours with *RB1* alterations, however these patients had a worse survival (median OS 1.4 years) compared to the HRP group with no *RB1* alteration (median OS 2.4 years).

Examination of genomic features revealed relatively similar patterns within *BRCA1*-HRD and *BRCA2*-HRD groups, although there were a few discriminatory features identified between those with and without *RB1* alterations (Supplementary Figs. S5 and S6). For example, the *BRCA1*-associated rearrangement signature Ovary_G^47^ was more enriched in *BRCA1*-HRD tumours with *RB1* alterations compared to those without (*P* = 0.039). Among *BRCA2*-HRD tumours, the mutational signatures DBS6 (unknown etiology) and SBS3 (associated with HRD)^48^ were higher in *RB1*-altered tumours compared to non-altered tumours, although this was not significant (*P* = 0.082 and *P* = 0.1 respectively). Concordantly, the average *BRCA1*-type and *BRCA2*-type CHORD scores^40^ were highest in *BRCA1*- and *BRCA2*-HRD tumours with *RB1* alterations respectively, indicating a higher probability of HRD. As described previously^49^, *CCNE1* gene amplifications were absent in tumours with both HRD and *RB1* alterations (*P* = 0.0006; Supplementary Fig. S7).

We hypothesised that tumours with combined HRD and *RB1* loss may have unique transcriptional profiles. To explore this, we compared gene expression profiles between each HRD/*RB1* group and the reference set of tumours that were HRP and *RB1* wild-type (Supplementary Table S7, Supplementary Fig. S8). There was significant enrichment of MSigDB hallmark gene sets among genes differentially expressed in *BRCA1*-HRD tumours with *RB1* alterations, the most prominent being interferon gamma response (up), interferon alpha response (up), oxidative phosphorylation (up), and E2F targets (up; adjusted *P* < 0.0001; Fig. 4A). The differentially expressed genes identified between *BRCA2*-HRD / *RB1* altered tumours and the reference set were significantly enriched for the MSigDB hallmark gene sets: E2F targets (up), epithelial mesenchymal transition (down), G2M checkpoint (up), and TNF alpha signalling via NF-kB (up; adjusted *P* < 0.0001).

**Figure 4.**
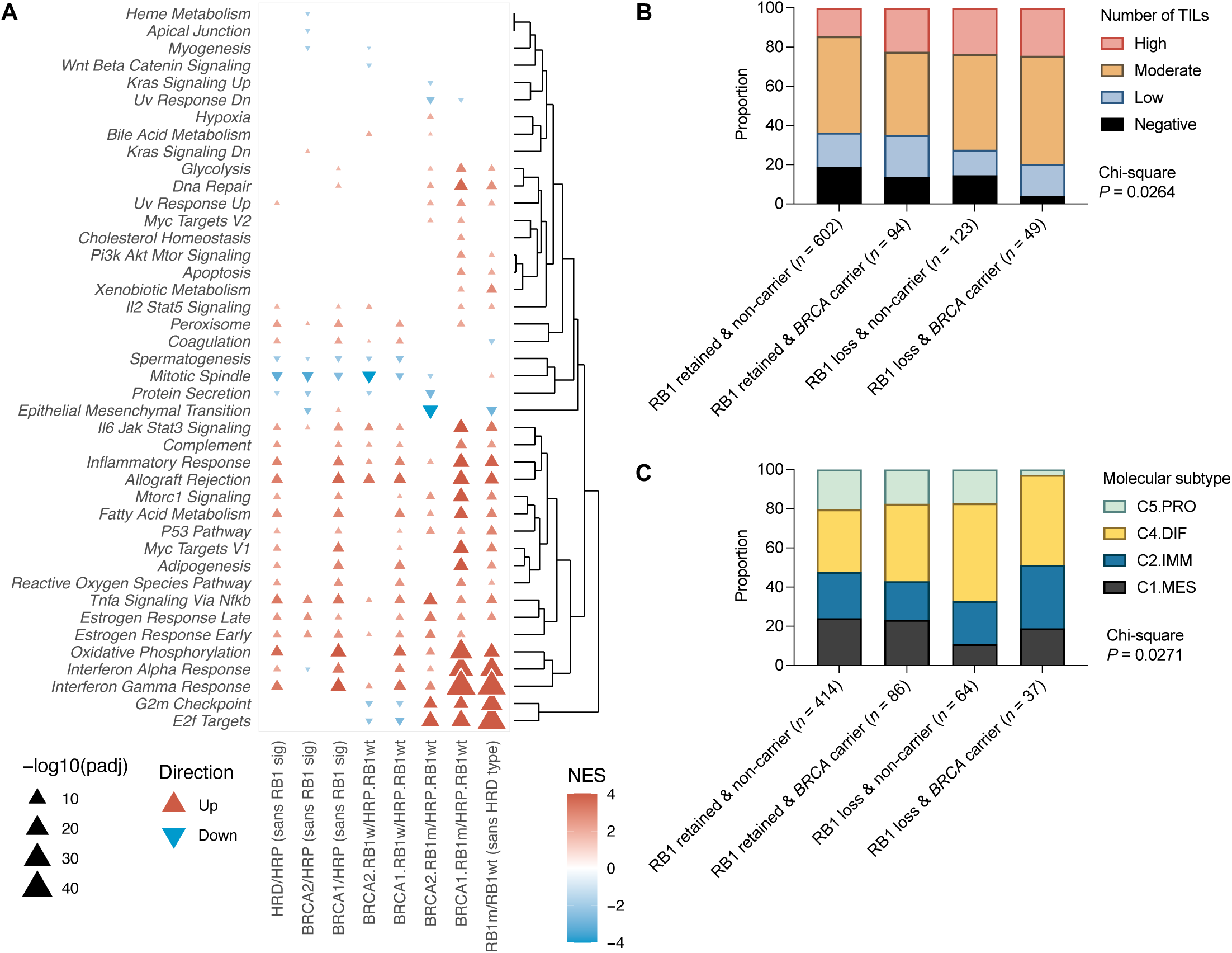
Characterisation of HGSC with co-loss of RB1 and *BRCA*. (A) Gene set enrichment analysis indicating up- and downregulated pathways in tumours according to *BRCA* and *RB1* status. HRP, homologous recombination proficient; HRD, homologous recombination deficient; RB1wt, *RB1* wild-type; RB1m, *RB1* altered. (B) Proportion of tumour infiltrating lymphocytes (TILs) in HGSC tumours grouped by RB1 expression and *BRCA* germline mutation status (Chi-square *P* value is indicated). (C) Proportion of tumours classified as each HGSC molecular subtype^12^ grouped by RB1 expression and *BRCA* germline mutation status (Chi-square *P* value is indicated; C5.PRO, C5/proliferative subtype; C4.DIF, C4/differentiated subtype; C2.IMM, C2/immunoreactive subtype; C1.MES, C1/mesenchymal subtype).

Since enhanced tumour cell proliferation has been associated with long-term survival in HGSC^7,26^, and loss of RB1 might accelerate proliferation^31^, we evaluated the expression of proliferation markers across the *RB1* and *BRCA* subgroups. *BRCA1*-HRD tumours with *RB1* alterations had significantly higher mRNA levels of the cell proliferation related genes *PCNA* (proliferating cell nuclear antigen) and *MCM3* (minichromosome maintenance complex component 3) compared to *BRCA1*-HRD tumours without *RB1* alterations (*P* < 0.0001, Supplementary Fig. S6). However, there were no significant differences in the proportion of Ki-67 positive cancer cell nuclei (*P* = 0.3297) across the subgroups (Supplementary Fig. S6), which was previously quantified by immunohistochemistry^7^ in a subset of primary tumours (*n* = 59).

### Germline BRCA mutation carriers with somatic loss of RB1 tumour expression show elevated immune activity

Having observed that HGSC with combined RB1 loss and HRD have enrichment of transcriptional signatures associated with an enhanced immune response, we accessed existing immunohistochemical data^39^ to determine the prevalence of CD8+ TILs in HGSC samples that also had RB1 protein expression and *BRCA* germline mutation status (*n* = 868). *BRCA* carriers with RB1 loss had a significantly higher proportion of tumours (79.6%) with moderate and high densities of CD8+ TILs, compared to *BRCA* carriers with retained RB1 (64.9%), non-carriers with RB1 loss (72.4%) and non-carriers with retained RB1 (63.6%, *P* = 0.0264; Fig. 4B). Tumours with complete absence of CD8+ TILs were the least frequent in *BRCA* carriers with RB1 loss (4.1%) compared to the other groups (13.8 % of *BRCA* carriers with retained RB1 tumour expression, 14.6% of non-carriers with RB1 tumour loss, 18.8% of non-carriers with retained RB1 tumour expression).

Gene expression-based molecular subtypes^12,38^ also differed by RB1 and *BRCA* status (*P* = 0.0271, *n* = 601; Fig. 4C). As expected, there was enrichment for the C2/immunoreactive subtype, a subtype characterised by the presence of intratumoural CD8+ T cells and good survival, in germline *BRCA* carriers with RB1 loss (32.4%) compared to the other subgroups (between 19.8% and 23.4%). Additionally, tumours with RB1 loss were enriched for the C4/differentiated molecular subtype, a subtype characterised by cytokine expression and good survival, regardless of *BRCA* status (45.9% in *BRCA* carriers with RB1 loss, 50.0% in non-carriers with RB1 loss, 39.5% in *BRCA* carriers with retained RB1, 32.1% of non-carriers with retained RB1). *BRCA* carriers with RB1 loss also had the lowest proportion of the C5/proliferative molecular subtype (2.7% versus 17.2% to 20.3% in the other groups), a subtype associated with diminished immune cell infiltration and poor survival^12,19^.

## DISCUSSION

Identifying the determinants of long-term patient survival, particularly in cancers with a generally unfavourable prognosis such as HGSC, may reveal novel therapeutic targets and inform personalised treatment strategies^8^. Improved survival associated with RB1 loss has been described previously in HGSC^7,34,35,50^ but the underlying factors contributing to this survival benefit have not been studied to date. We assessed tumour samples from a cohort of more than 7,000 women with ovarian cancer, including a subset with high resolution genomic data, to understand how RB1 loss may impact on therapeutic response and patient survival.

Alteration of the RB1 pathway is a frequent event in tumourigenesis, including loss of regulators such as p16, activation of D- and E-type cyclins and their associated cyclin dependent kinases, and loss of RB1 itself (reviewed in ^51^). Our study showed that RB1 loss is associated with longer survival in patients with advanced stage HGSC, but by contrast, loss of RB1 in ENOC was associated with a shorter survival, particularly in combination with p53 mutation. Similar to ENOC, in endocrine-driven breast and prostate cancer, RB1 loss is associated with poorer survival: early co-loss of *BRCA2* and *RB1* is associated with an aggressive, castration-resistant prostate cancer subtype (CRPC) characterised by epithelial-to-mesenchymal transition and shorter survival^29^. RB1 loss facilitates lineage plasticity and, with p53-comutation, leads to an androgen-independent phenotype^52,53^ and consequently resistance to anti-androgen therapy. In estrogen-receptor (ER) positive breast cancer, CDK4/6 inhibitor resistance is associated with RB1 loss and cyclin E2 activation^54,55^.

Triple negative breast cancer (TNBC) provides an important contrast to the findings for RB1 loss in ER-positive breast cancer. In TNBC, RB1 loss is most common in the basal-like subtype, where *BRCA1* mutation and promoter hypermethylation is associated with frequent *RB1* gene disruption and RB1 loss^28^. RB1 loss alone, as well as co-occurrence with *BRCA1* promoter hypermethylation, is associated with a favourable chemotherapy response and outcome^27,56–58^. Notably, TNBC and HGSC are more similar than the cancers that they are grouped with anatomically, sharing gene expression patterns, genetic drivers including *BRCA1* and *BRCA2*, ubiquitous loss of *TP53*, extensive copy number variation, and susceptibility to platinum-based chemotherapy^59,60^. Taken together, the relationship between RB1 loss and patient survival appears to be dependent on cancer type and molecular context^61^.

Some, but not all TNBC and early metastatic prostate cancers are associated with germline variants in *BRCA1*, *BRCA2* and other genes involved in HR DNA repair. However, previous tumour studies of RB1 expression have not also defined the HRD status of individual samples. A strength of this study was the known *BRCA* germline status of 1134 of the HGSC patients for which we also had RB1 protein expression, and this revealed the strong association of co-mutation in either *BRCA1* or *BRCA2* and *RB1* with survival. In addition to germline mutations in *BRCA1* or *BRCA2*, germline or somatic mutations, and promoter methylation of other genes involved in HR DNA repair, such as *RAD51C*, can result in a similar molecular phenotype, characterised by distinct genomic scarring^26^. Using whole-genome sequence data, we determined the likely tumour HRD status in a subset of 126 tumours using an algorithm that recognises genomic scarring associated with HRD (Fig. 3A), rather than simply designating *BRCA* mutation status, which does not account for all mechanisms of HR repair inactivation. Although the number of samples with RB1 loss and HR proficiency was small, the very poor outcome we observed with this group indicated that for RB1 to impart a survival benefit in HGSC, it must occur in an HRD background. Validation of this finding in a larger cohort may further inform how RB1 loss could favourably influence survival in certain histological and molecular contexts.

We have previously noted that enhanced proliferation in HGSC is associated with long-term survival^7,26^ and it is reasonable to suggest that RB1 loss may be imparting an effect through deregulating the cell cycle. However, data on the effect of RB1 loss on proliferation in HGSC tumours and cancer cell lines is inconsistent. *RB1* knockout in our HGSC cell lines did not cause cell cycle alterations in the absence of treatment, and despite differences in proliferative markers at the mRNA level, there was no significant difference in the proportion of Ki-67 positive nuclei between tumours with or without RB1 protein expression. In a recent OTTA study, Ki-67 expression was not associated with survival in HGSC; however, there was strong correlation between loss of RB1 and the proliferative marker MCM3^62^, which may provide a more accurate measure of tumour cell proliferation than Ki-67^63^.

In addition to its role in driving progression through the G1 stage of the cell cycle, RB1 has non-canonical functions. RB1 has been shown to participate in HR DNA repair through interactions with BRG1 and ATM^64^. A recent pan-cancer study^65^ found that combined loss of *TP53* and *RB1* was associated with a particularly high genome-wide loss-of-heterozygosity score, one of the key elements of genomic scarring associated with HRD. In our whole-genome analysis, HGSC tumours with dual loss of HRD and *RB1* did not exhibit overall higher mutation burden; however, we did observe elevated levels of mutational signatures associated with HRD, which may be evidence of compounding DNA repair defects. It remains possible that the combined inactivation of RB1 and HR genes contribute to enhanced chemotherapy response and/or an impaired ability for tumour cells to develop therapy resistance.

When we evaluated a set of patient derived HGSC lines, those with germline *BRCA1* mutation and *RB1* alteration were most sensitive to cisplatin and olaparib. Knockout of *RB1* in the AOCS 7.2 cell line which had a pre-existing *BRCA1* mutation, resulted in an increase in chemosensitivity, consistent with the notion that co-mutation enhances chemotherapy response^7^. Unfortunately, despite considerable efforts, we were unable to generate a larger series of isogenically matched cell lines with combinations of conditional knockouts of *RB1* and *BRCA1* as all surviving clones retained at least one *BRCA1* allele. *BRCA1* loss is embryonic lethal and engineered loss in cell lines has been reported as lethal elsewhere including in the human haploid cell line, HAP1^46^.

Our data provides evidence of an enhanced immunogenicity in HGSC with RB1 loss, with higher CD8+ TIL counts and upregulated expression of IFN-γ signalling pathways. RB1 has been shown to inhibit innate IFN-β production in immunocompetent mice^66^ and RB1 deficiency triggered an increased IFN-β and IFN-α secretion. Co-mutation of *RB1* and *TP53* was recently found to be associated with an enhanced response to the immune checkpoint inhibitor atezolizumab in metastatic urothelial bladder cancer^67^. Similarly, a case report described a complete response to atezolizumab in heavily pre-treated, RB1-negative TNBC^68^. This generates the hypothesis that RB1 loss could predict response to such therapies in HGSC, since this tumour type ubiquitously harbours *TP53* mutations^69^. However, a recent biomarker study in ovarian cancer patients treated with atezolizumab or placebo and standard chemotherapy found that deleterious mutations in *RB1* were prognostic for a better PFS, regardless of the addition of atezolizumab^70^. While it appears RB1 loss alone may not be predictive of response to the PD-L1 inhibitor atezolizumab, response rates to PD-1/PD-L1 pathway checkpoint inhibitors are generally quite low in HGSC, with the best objective response rates between 8% and 15%^71^. Our study has identified a subset of patients with combined RB1 and *BRCA* inactivation who demonstrate exceptional immune responses and may provide clues for the development of new immunotherapeutic strategies for HGSC that extend beyond targeting PD-L1/PD-1.

Our work highlights the importance of RB1 loss to treatment response and survival and focuses attention on other therapeutic opportunities in this subset of HGSC patients. Approximately 20 percent of HGSC patients have somatic loss of *RB1* assessed using genomic data^3,26^, a figure that is consistent with the immunohistochemical results obtained in the large patient cohort described here. Both approaches indicate that RB1 loss is generally clonal, enhancing its value as a therapeutic target if selective inhibitors can be identified. Casein kinase 2 (CK2) inhibitors have been reported to enhance the sensitivity of *RB1*-deficient TNBC and HGSC cells to carboplatin and niraparib^72^. In addition, Aurora kinase A and B inhibition is synthetically lethal in combination with RB1 loss in breast and lung cancer cells^73–75^. Irrespective of HRD status, *RB1* mutations correlate with sensitivity to WEE1 inhibition in *TP53* mutant TNBC and HGSC patient-derived xenografts^76^, indicating additional treatment options that exploit RB1 inactivation in these tumours. In this study, the *BRCA1*-mutant cell line AOCS7.2 with induced *RB1* knockout was more sensitive to olaparib suggesting that RB1 loss may also predict responses to PARP inhibitors in HGSC. RB1 staining of tumour tissue by IHC is a relatively low-cost pathology-based assay that could be used in prospective studies to test whether RB1 expression is predictive of responses to PARP inhibitors, either alone or in combination with approved HRD tests.

## Data Availability

All data produced in the present study are available upon reasonable request to the authors.

## ACKNOWLEDGMENTS

We thank J. Beach and L. Bowes for their contributions to the study. This work was supported by the National Health and Medical Research Council (NHMRC) of Australia (1186505 to DWG; 1092856, 1117044 and 2008781 to DDLB; 2009840 to SJR), the National Institutes of Health (NIH) / National Cancer Institute (R01CA172404 to SJR, P50 CA136393 to SHK) and the U.S. Army Medical Research and Materiel Command Ovarian Cancer Research Program (Award No. W81XWH-16-2-0010 and W81XWH-21-1-0401). DWG is supported by a Victorian Cancer Agency / Ovarian Cancer Australia Low-Survival Cancer Philanthropic Mid-Career Research Fellowship (MCRF22018). FAMS is supported by a Swiss National Foundation Early Postdoc Mobility Fellowship (P2BEP3-172246), a Swiss Cancer League grant BIL KFS-3942-08-2016 and a Prof. Max Cloëtta foundation grant. KIP is supported by a NHMRC CJ Martin Overseas Biomedical Fellowship (APP1111032). ELC is supported by a Victorian Cancer Agency Mid-Career Fellowship (MCRF21004). MW is supported by the European Research Council under the European Union’s Horizon 2020 Research and Innovation Programme grant agreement No 742432 (BRCA-ERC). KS is supported by the Swedish Cancer Foundation. MSA is funded through a Michael Smith Health Research BC Scholar Award (18274) and the Janet D. Cottrelle Foundation Scholars program managed by the BC Cancer Foundation.

BC’s Gynecological Cancer Research team (OVCARE) receives support through the BC Cancer Foundation and the VGH & UBC Hospitals Foundation. The Gynaecological Oncology Biobank at Westmead was funded by the NHMRC (ID310670, ID628903); the Cancer Institute NSW (12/RIG/1-17, 15/RIG/1-16); and acknowledges support from the Department of Gynaecological Oncology, Westmead Hospital, and the Sydney West Translational Cancer Research Centre (Cancer Institute NSW 15/TRC/1-01). The Women’s Cancer Research Program at Cedars-Sinai Medical Center (LAX) is supported by The National Center for Advancing Translational Sciences (NCATS) Grant UL1TR000124. The Study of Epidemiology and Risk Factors in Cancer Heredity (SEARCH) is funded by Cancer Research UK (C490/A10119 C490/A10124 C490/A16561) and the UK National Institute for Health Research Biomedical Research Centre at the University of Cambridge. The UKOPS study was funded by The Eve Appeal (The Oak Foundation) with contribution to authors’ salary through MRC core funding MC_UU_00004/01 and the National Institute for Health Research University College London Hospitals Biomedical Research Centre.

The investigators also acknowledge generous contributions from the Border Ovarian Cancer Awareness Group, the Peter MacCallum Cancer Foundation, the Graf Family Foundation, Wendy Taylor, Arthur Coombs and family, and the Piers K Fowler Fund. The contents of the published material are solely the responsibility of the authors and do not reflect the views of the NHMRC, NIH, and other funders.

## AUTHOR CONTRIBUTIONS

MK, SJR, DDLB and DWG conceived the study design. FAMS, KT, KP, JB and TH carried out experiments, and analysed and interpreted results along with TB, AP, DA, TZ, NSM, SF, AD, MK, SJR, DDLB and DWG. MK assessed and interpreted immunohistochemical scores. All authors contributed through recruitment and consenting of patients, collection and processing of biological samples, clinical care, abstraction and curation of clinical data and maintenance of follow-up. DDLB and DWG supervised the study and together with FAMS and KT wrote the manuscript. All authors contributed to writing, review and revision of the manuscript and approved the final submitted version.

---

**Supplementary Figure S1.**
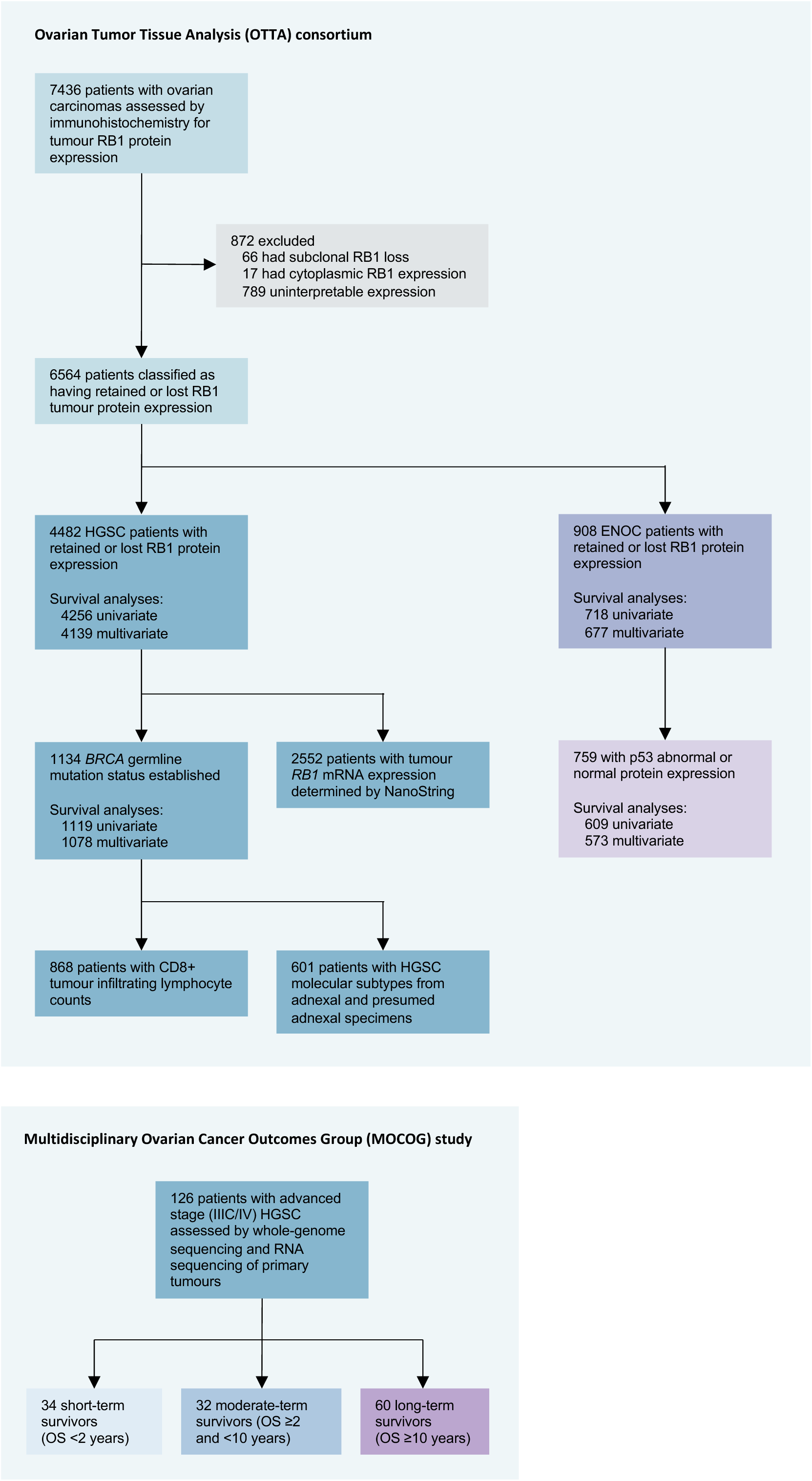
Patients and tumour samples analysed in this study. Number of patients included in each molecular analysis. HGSC, tubo-ovarian high-grade serous ovarian carcinoma; ENOC, endometrioid ovarian carcinoma; OS, overall survival.

**Supplementary Figure S2.**
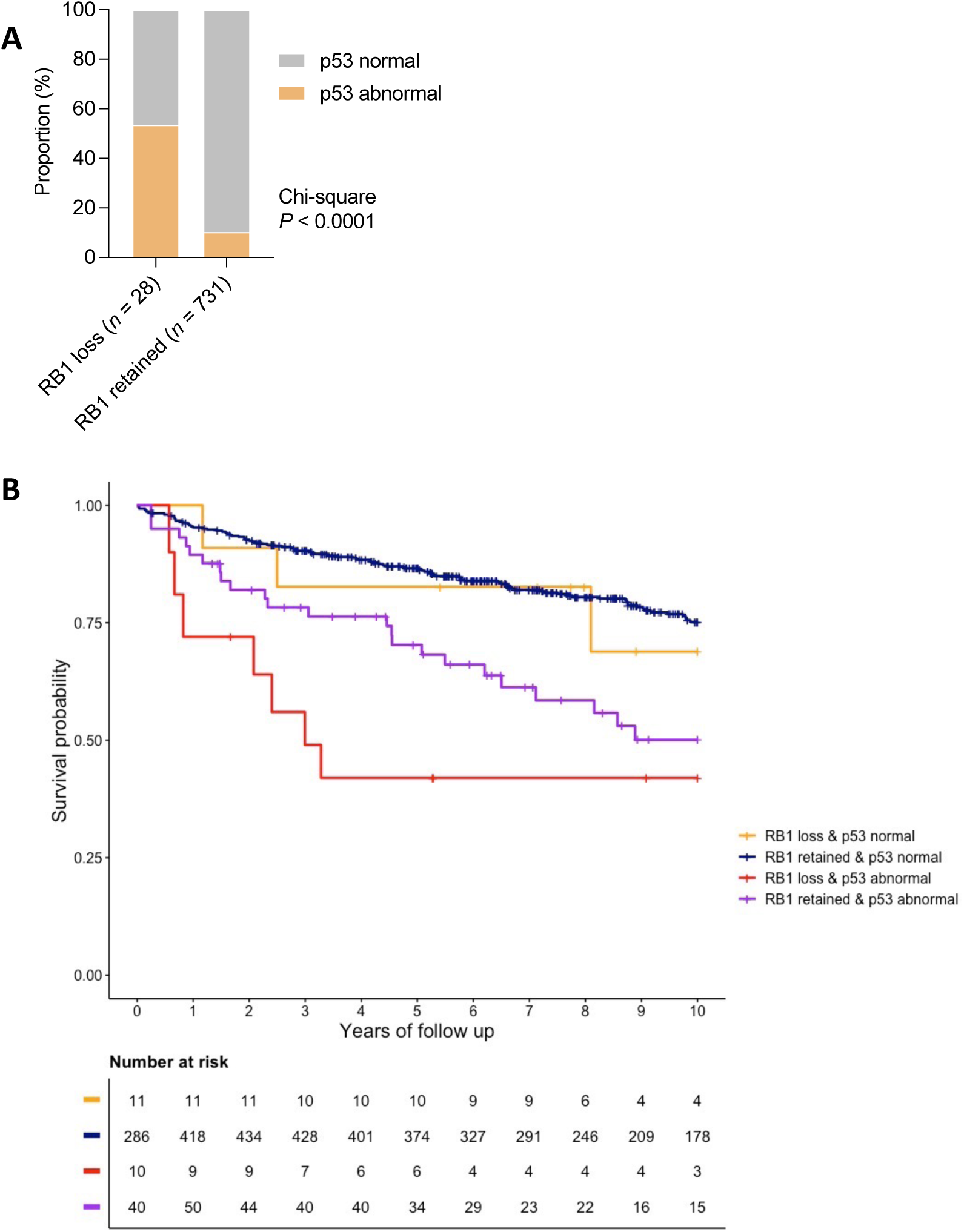
Combined p53 and RB1 protein expression in ENOC. (A) Correlation between RB1 and p53 tumour expression in patients with endometrioid ovarian carcinoma (ENOC). Chi-square *P* value is reported. (B) Kaplan-Meier estimates of overall survival in patients with ENOC by combined RB1 and p53 tumour expression status.

**Supplementary Figure S3.**
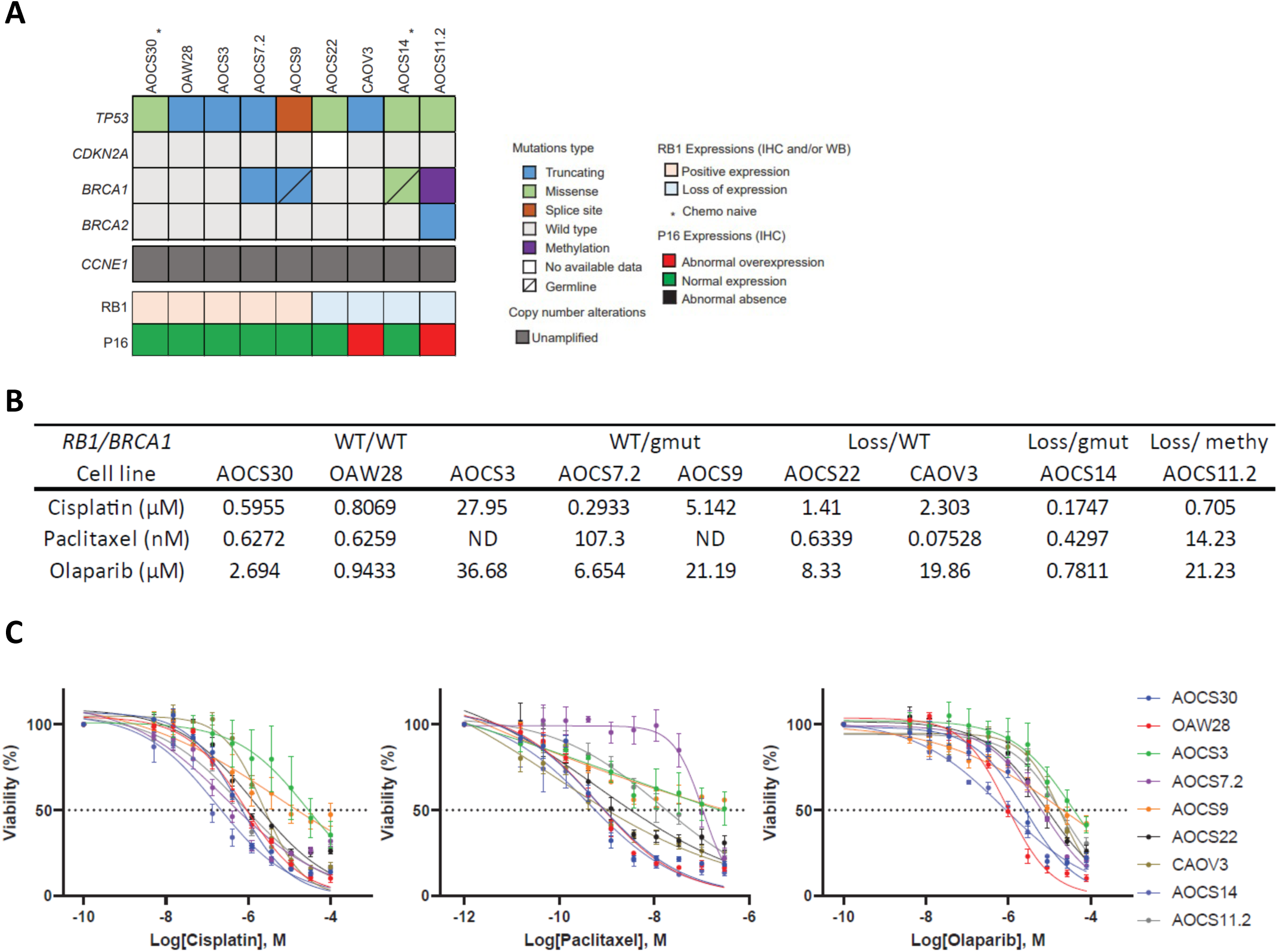
Drug sensitivity in HGSC cell lines with innate *RB1* and/or *BRCA1* alterations. (A) Summary of the molecular features of innate HGSC cell models, including mutations in key genes (*TP53*, *CDKN2A*, *BRCA1*, *BRCA2*), copy number alterations in *CCNE1*, and protein expression of RB1 and p16. (B) IC50 of high grade serous ovarian cancer cell lines after treatment with cisplatin (72 hours), paclitaxel (72 hours), or olaparib (120 hours). ND, Not determined. (C) Viability of high-grade serous ovarian cancer cell lines after treatment with cisplatin (72 hours), paclitaxel (72 hours), or olaparib (120 hours). Data are expressed as mean (*n* = 3 replicates) ± standard error of the mean (SEM). For some points, error bars are shorter than the height of the symbol and are not visible.

**Supplementary Figure S4.**
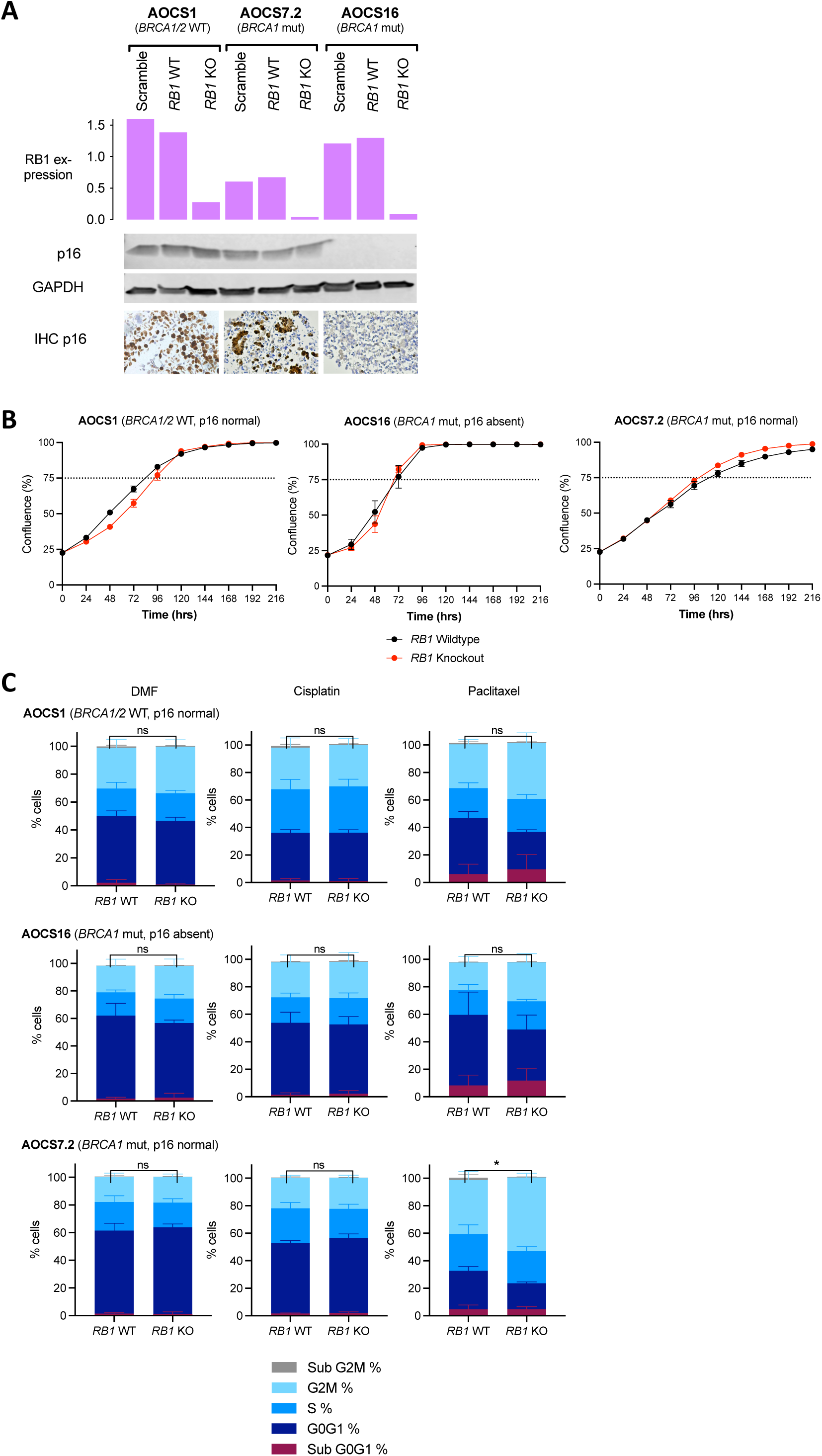
Cell proliferation and cell cycle distribution of HGSC cell lines with *RB1* knockout. (A) CRISPR/Cas9 knockout of *RB1* in 3 patient-derived ovarian cancer cell lines with different *BRCA1/2* and p16 backgrounds. The bar graph indicates *RB1* mRNA expression levels determined by RT-PCR (*n* = 3) in single-cell clones confirming *RB1* wildtype (WT) and knockout (KO) compared to heterozygous colonies without gene editing (Scramble). Representative Western Blots show p16 protein levels compared to GAPDH loading controls in each cell line and clone. Images of p16 IHC in AOCS parental cell lines are included confirming the respective p16 status. (B) Proliferative capacity of 3 patient-derived HGSC cell lines (*RB1* wild-type, WT and *RB1* knockout, KO clones) measured by IncuCyte Zoom live-cell imaging. Data represent mean ± SEM confluency after 20-25% starting confluency from three to six independent experiments. Dashed line denotes 75% confluency. (C) Cell cycle distribution following *RB1* CRISPR knockout. Proportion of cells in G0G1, S or G2/M phase 24 hours after treatment with DMF, cisplatin or paclitaxel at half the IC50 determined per cell line and drug, analysed by flow cytometry. Mean proportion ± SEM of three independently performed experiments are shown. Distribution was compared between *RB1* WT and KO clones using unpaired t test (ns, not significant; \**P* < 0.05).

**Supplementary Figure S5.**
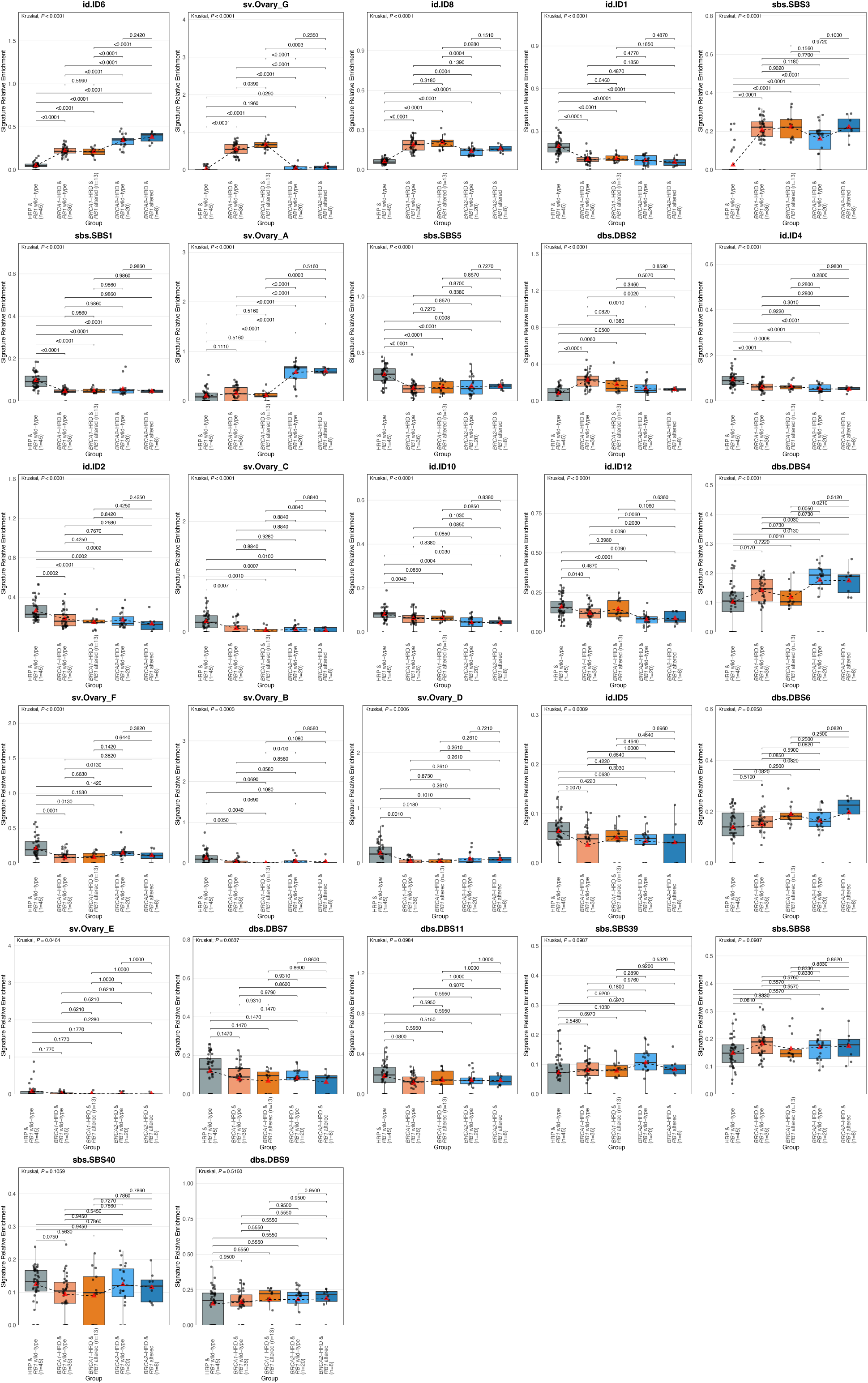
Mutational signatures in homologous recombination deficiency and *RB1* subgroups. Boxplots show the relative proportion (y-axis) of genome-wide mutational signatures^26^ according to homologous recombination deficiency (HRD) and *RB1* status. Boxes show the interquartile range (25-75^th^ percentiles), central lines indicate the median, dots represent each sample, whiskers show the smallest and largest values within 1.5 times the interquartile range, red triangles indicate the mean, and dotted lines join the mean of each subgroup to visualise the trend. The Kruskal–Wallis test *P* values displayed are Benjamini-Hochberg adjusted and the signatures are ordered by their significance. Pair-wise Mann-Whitney-Wilcoxon test adjusted *P* values are also reported. HRP, homologous recombination proficient.

**Supplementary Figure S6.**
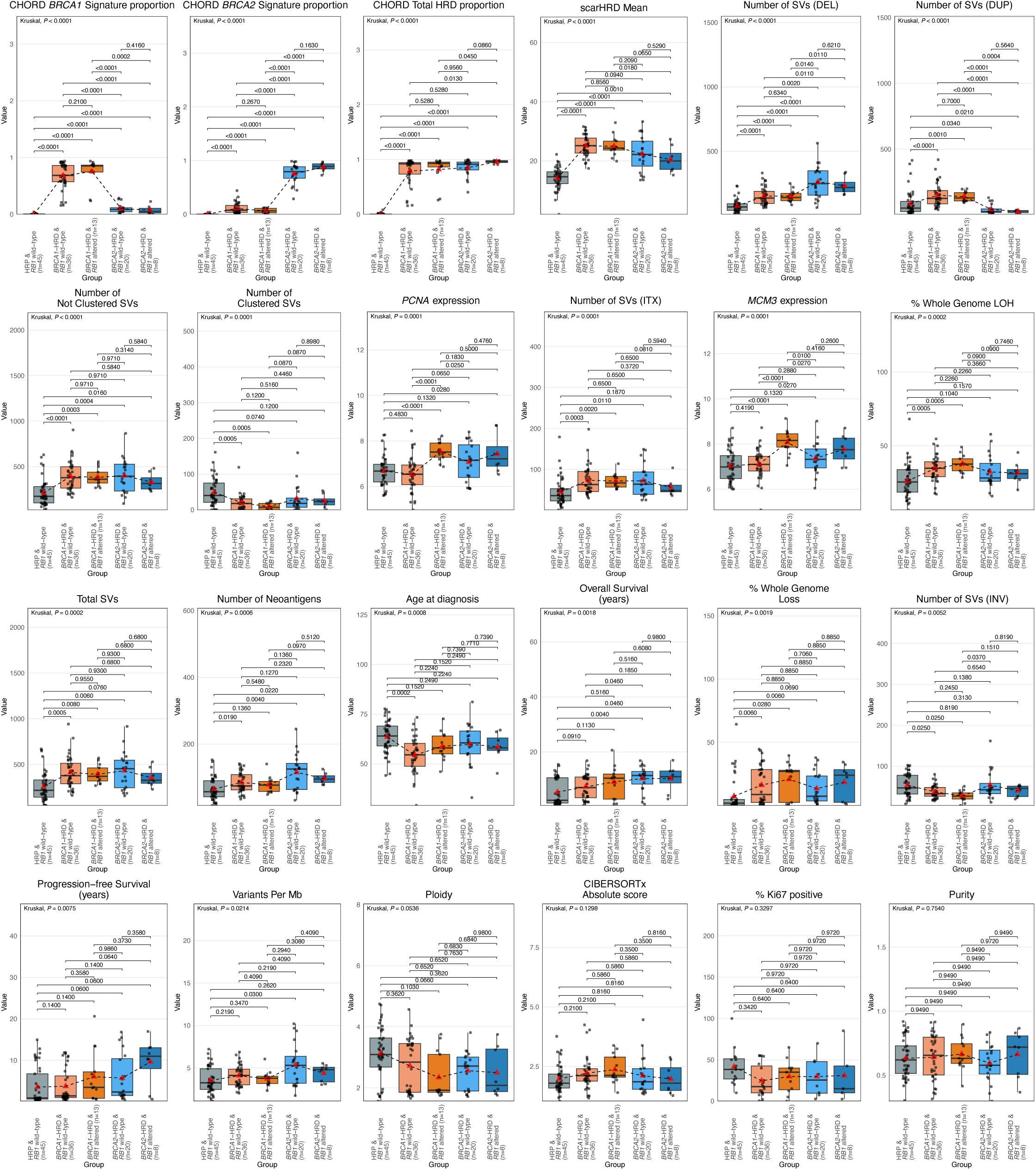
Genomic and clinical characteristics by combined homologous recombination deficiency and *RB1* status. Boxplots show numerical clinical and genomic features (y-axis) according to homologous recombination deficiency (HRD) and *RB1* status. Boxes show the interquartile range (25-75^th^ percentiles), central lines indicate the median, dots represent each sample, whiskers show the smallest and largest values within 1.5 times the interquartile range, red triangles indicate the mean, and dotted lines join the mean of each subgroup to visualise the trend. The Kruskal– Wallis test *P* values displayed are Benjamini-Hochberg adjusted and the features are ordered by their significance. Pair-wise Mann-Whitney-Wilcoxon test adjusted *P* values are also reported. Features include *BRCA1*- and *BRCA2*-type CHORD (Classifier of HOmologous Recombination Deficiency) scores; mean HRD scores (scarHRD); absolute numbers of structural variants (SVs), including deletions (DEL), duplications (DUP), intrachromosomal rearrangements (ITX), and inversions (INV); relative expression levels of *PCNA* and *MCM3*; proportion of whole-genome loss-of-heterozygosity (LOH); number of predicted neoantigens and variants per megabase (Mb); age of patients at diagnosis; progression-free and overall survival; cancer cell purity and ploidy; absolute CIBERSORTx scores; proportion of Ki-67 positive tumour cells were available for *n* = 59 primary tumours as previously measured by immunohistochemistry^7^. HRP, homologous recombination proficient.

**Supplementary Figure S7.**
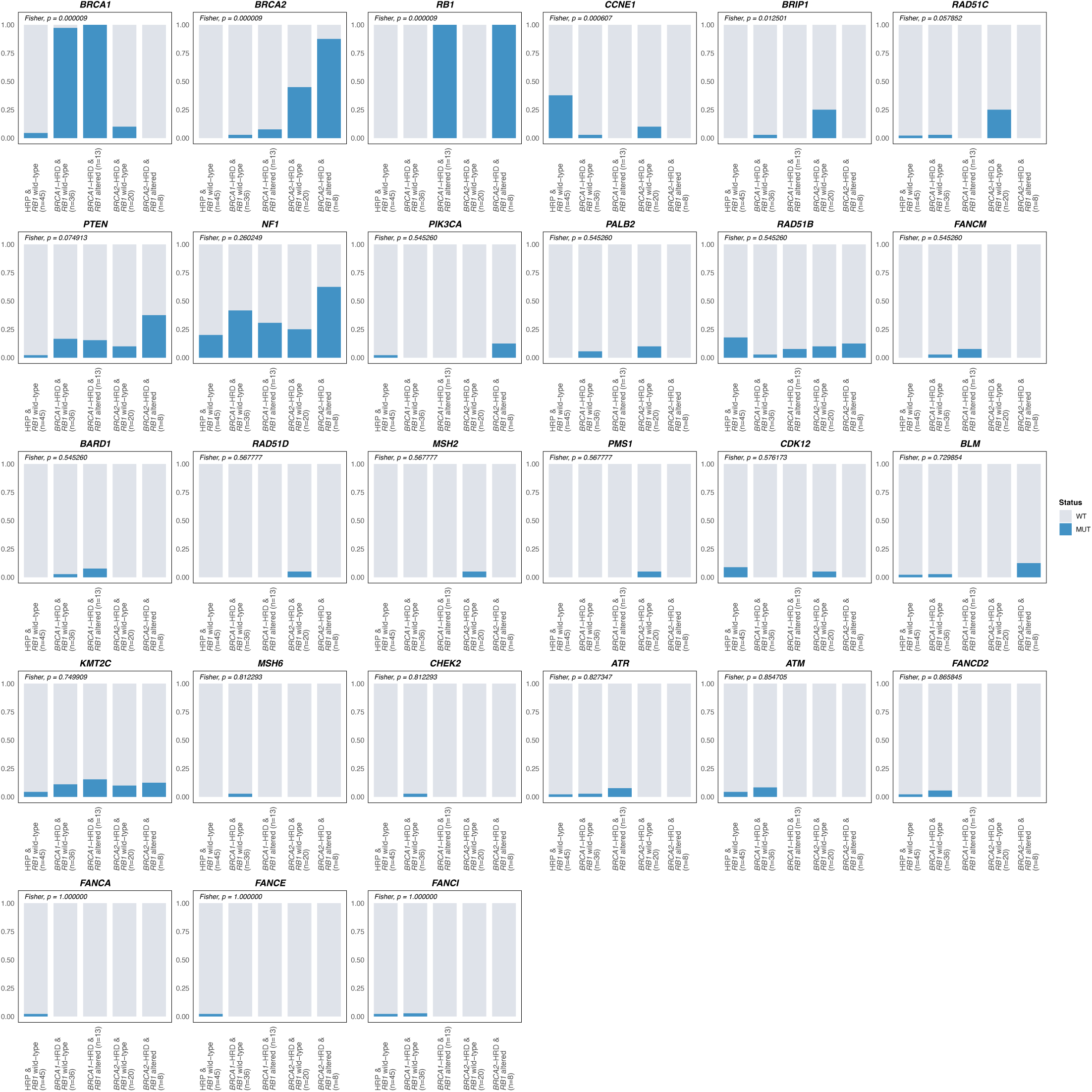
**Gene alterations across *BRCA* and *RB1* altered subgroups.** Proportion of tumours with alterations in genes of interest for each subgroup. WT, wild-type; MUT, mutation; HRP, homologous recombination proficient. Genes are ordered by significance using Fisher’s exact test; Benjamini-Hochberg adjusted *P* values are reported.

**Supplementary Figure S8.**
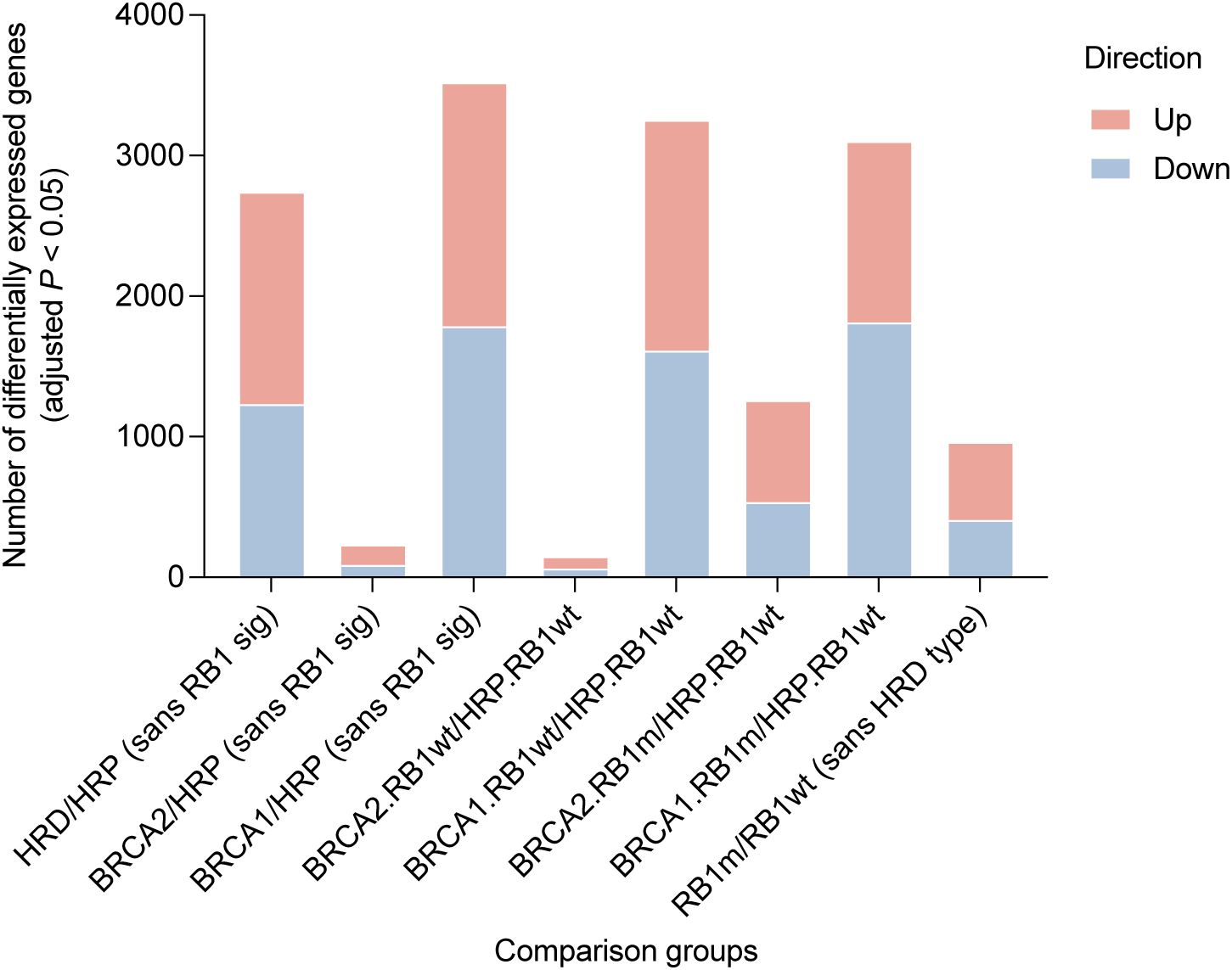
Differentially expressed genes. Bars indicate the number of differentially expressed genes (Benjamini-Hochberg adjusted *P* value < 0.05) between HGSC tumours grouped by HRD and/or *RB1* status as shown. Differential gene expression analysis was performed using DESeq2 to determine fold change of gene expression between groups (see Supplementary Table 7 for full DESeq2 results). HRP, homologous recombination proficient; HRD, homologous recombination deficient; RB1wt, *RB1* wild-type; RB1m, *RB1* altered.

