## Supplementary Material for "Concurrent RB1 loss and *BRCA*-deficiency predicts enhanced immunological response and long-term survival in tubo-ovarian high-grade serous carcinoma"

***Immunohistochemical (IHC) staining and analysis of RB1 protein expression***

Formalin-fixed paraffin-embedded (FFPE) samples on slides were subjected to heat induced antigen retrieval with Target Retrieval Solution (TRS) high on the DAKO Omnis platform (Agilent Technologies, Santa Clara, CA, USA), then incubated with anti-RB1 (Retinoblastoma Gene Protein) mouse monoclonal antibody (Leica, Clone 13A10, Novocastra: #NCL-L-RB-358) at a 1:100 dilution. Staining was visualised using 3,3'-diaminobenzidine (DAB). Scoring was conducted by a pathologist; samples were scored as either 0 (absent RB1 expression with RB1 expression present in normal cell serving as internal control), 1 (RB1 present), 2 (subclonal loss of RB1 expression), 3 (cytoplasmic staining) or 8 (RB1 absent but lacking adjacent internal control). Representative images of RB1 expression patterns in tumour tissue are shown below.

| | |
|---|---|
| 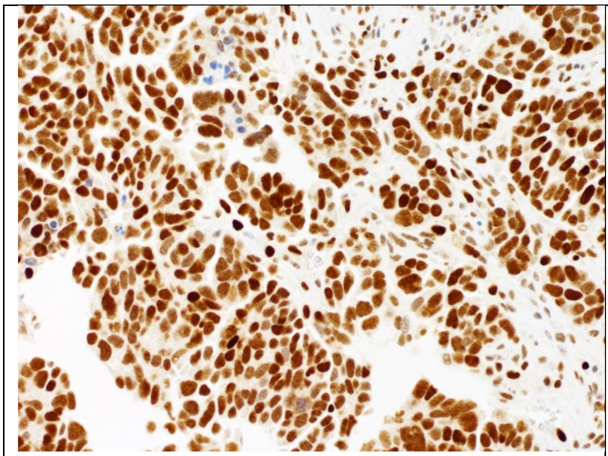 | 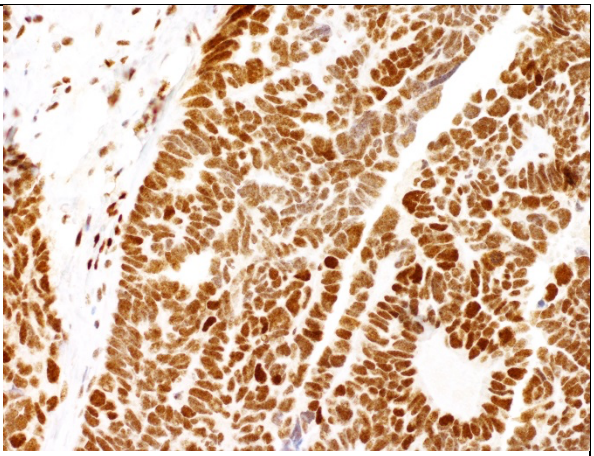 |
| Score 1 = present | Score 1 = present |

| | |
|---|---|
| 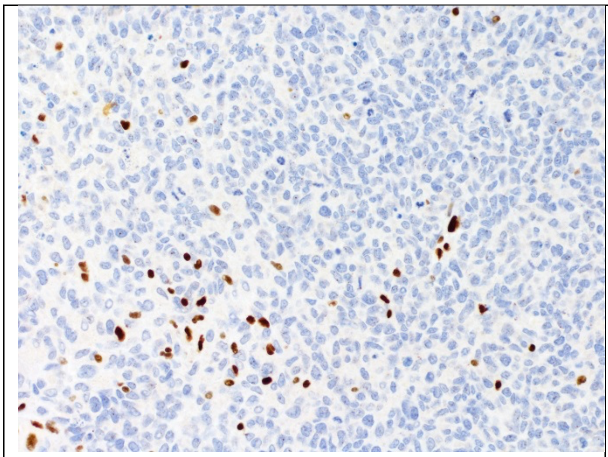 | 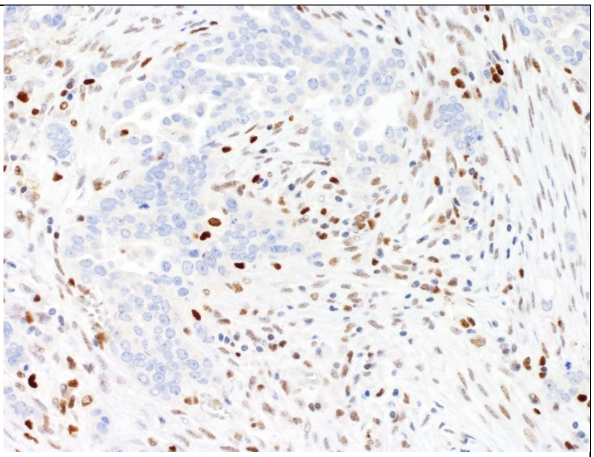 |
| Score 0 = absent | Score 0 = absent |
| 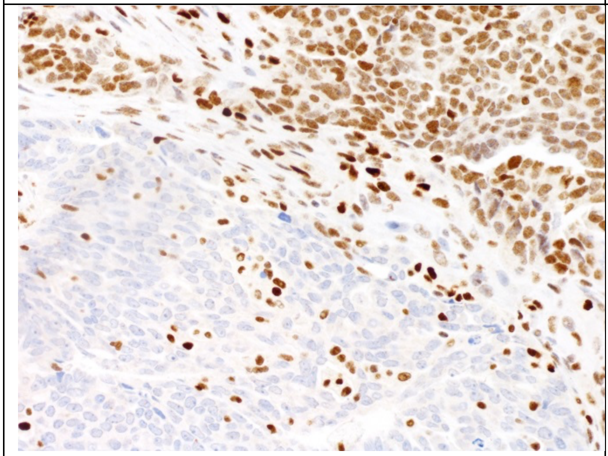 | 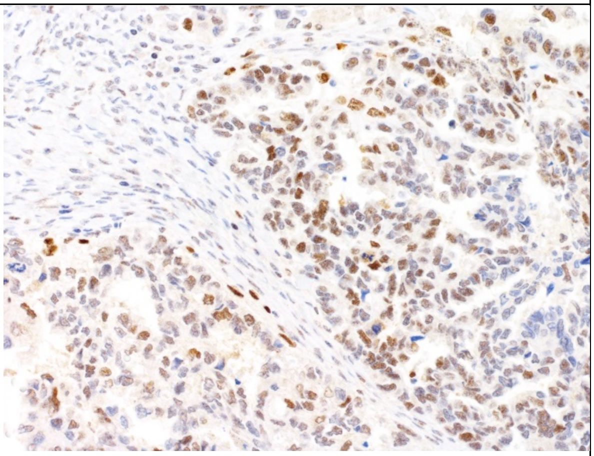 |
| Score 2 = subclonal loss | Score 1 = present (although heterogeneously reduced) |
| 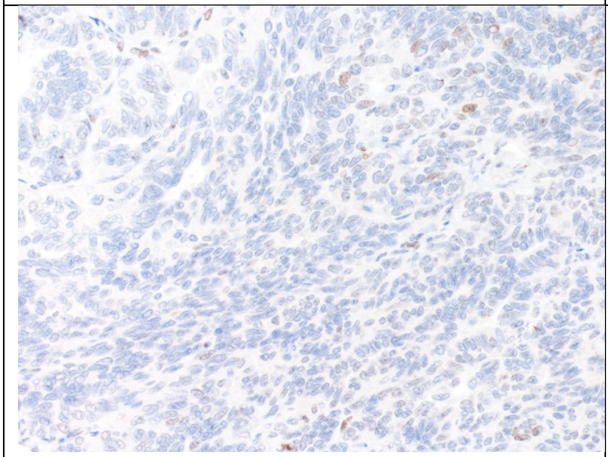 | |
| Score 8 = no staining and lack of internal control | |

### *Differential gene expression analysis*

Primary tubo-ovarian high-grade serous carcinoma (HGSC) tumour samples were grouped according to *RB1* alterations and homologous recombination deficiency (HRD) status, as assessed previously using whole-genome sequencing[1] and the CHORD (Classifier of Homologous Recombination Deficiency) method[2]. Matched RNA sequencing data was previously processed into gene expression counts as part of the prior Multidisciplinary Ovarian Cancer Outcomes Group (MOCOG) study[1]. Differentially expressed protein coding genes were identified between sample groups of interest using DESeq2[3] (v1.26.0), with batch effects accounted for in the model. In addition to characterising the transcriptional profiles of tumours with *RB1* alterations and concomitant *BRCA1*- or *BRCA2*-type HRD relative to tumours with no alterations, DESeq2 was also used to evaluate alteration-specific transcriptional profiles by incorporating given alterations into the model to remove their signal (each comparison is shown in Supplementary Table 7).

The R package FGSEA[4] (fast gene set enrichment analysis; v1.15.1) was used to perform gene set enrichment analyses across comparison groups. Gene level Benjamini-Hochberg adjusted *P* values obtained from DESeq2 were transformed to signed *P* values by converting them to a negative log10 value and applying the sign of the fold change. The signed *P* values were pre-sorted and fed into FGSEA via its function fgseaMultilevel (minSize=15, maxSize = 500, gseaParam = 0, eps = 0) to generate enrichment scores and adjusted *P* values using the MSigDB[5] Hallmark gene sets (v7.4).

### *Cell line growth conditions*

Cell lines were maintained in a humidified incubator at 37°C and 5% $CO_2$. All cell lines (aside from JHOS2) were cultured in RPMI 1640 (GIBCO, Carlsbad, CA, USA) supplemented with 10% fetal bovine serum (FBS, Cytiva) and 1% penicillin-streptomycin-glutamine (GIBCO).

JHOS2 was cultured in DMEM/F12 (GIBCO) with 15 mM HEPES, 10% FBS, 1X MEM non-essential amino acids (GIBCO, 100X) and 1% penicillin-streptomycin (GIBCO).

***Molecular characterisation of cell lines***

Complete cell line characterisation details can be found in Supplementary Tables S8 and S9. The mutation status of genes of interest in AOCS cell lines was determined by either whole-genome[1] or targeted sequencing[6,7] using established pipelines, and in commercial cell lines from published data[8] or The Cancer Cell Line Encyclopaedia in cBioPortal[9-11]. *BRCA* and *TP53* variants were classified as pathogenic if they were truncating (nonsense, splice site or frameshift) mutations resulting in early stop codons, or missense variants previously reported as pathogenic in ClinVar[12] or The TP53 Database (R20, July 2019, https://tp53.isb-cgc.org). *CCNE1* copy number in AOCS cell lines was analysed by qPCR in triplicate on LightCycler 480 (Roche) using SYBR Green PCR mix (Applied Biosystems) as described previously[13]. The expression status of RB1 and p16 was evaluated by Western blot (as below) and/or IHC. For IHC, FFPE cell line plugs were established by fixing approximately $6x10^7$ cells in 10% Neutral Buffered Formalin (NBF) overnight, transferring them into an agarose gel plug and embedding them in paraffin. Duplicate cores were taken from each cell line plug and assembled in a paraffin block in the fashion of a tissue microarray. Cell line microarrays were sectioned, stained with antibodies (RB1, BD Pharmingen, BD Biosciences, clone G3-245; p16, Roche Ventana, CINtec, clone E6H4) and scored blinded by a pathologist. RB1 was classified as either absent, present or uninterpretable; p16 was interpreted according to a 3-tier scoring system as normal patchy, abnormal absent or abnormal overexpressed.

***CRISPR-mediated gene knockout***

*RB1* was inactivated using CRISPR-Cas9[14] in cell lines with a pre-existing *BRCA1* mutation (AOCS7.2, AOCS16, JHOS2) and those that were *BRCA1/2* wildtype (AOCS1, OVCAR4). Briefly, lentiviral transduction was performed using the FgH1t vector co-expressing Cas9, mCherry, and GFP and a doxycycline inducible synthetic guide RNA (sgRNAs) targeting *RB1* exon 7 or exon 8 (Supplementary Table S10). After sorting for double positive cells (mCherry and GFP) by flow cytometry, expression of the sgRNA was induced with doxycycline (0.1µg/ml media, Sigma-Aldrich, D3072) for 96 hours, and single cells sorted into 96-well plates. Clones were expanded and *RB1* status confirmed by reduced/absent RB1 expression (Western blot, RT-qPCR) and Sanger sequencing of the targeted *RB1* exon. For control lines, *RB1* wild-type single cell colonies without a CRISPR-edit were used, as well as heterogeneous cell populations with transduced Cas9 and sgRNA of a scrambled DNA sequence[15] (Supplementary Table S10).

Dual gene knockout of *RB1* and *BRCA1* was performed in AOCS30 using nucleofection[16-18] rather than lentivirus transduction. *BRCA1*, *RB1*, and control sgRNA sequences (CRISPRevolution sgRNA EZ Kit, Synthego) were designed as previously described[19,20]. Cells ($5x10^5$) were trypsinised, washed twice with Phosphate Buffered Saline (PBS) and incubated with the RNP complex (Alt-R® S.p. Cas9 Nuclease purified Cas9 protein, Integrated DNA Technologies) for 10 minutes. Cell pellets were suspended with NucleofectorTM SE solution (Lonza Bioscience) and mixed with prepared Cas9/sgRNA RNP complex, which were transferred into the NucleocuvetteTM vessels (Lonza Bioscience). Nucleofection was conducted with CL-120 Program in 4D-Nucleofector X unit (Lonza Bioscience). Pre-warmed medium was added to cells and incubated for 10 minutes in a humidified 37°C incubator with 5% $CO_2$. Cells were transferred into 6-well plates and cultured. Each cell line (AOCS30 NT, AOCS30 *BRCA1*KO, AOCS30 *RB1*KO, AOCS30 *RB1BRCA1*KO) was passaged two times to expand following nucleofection, passed through a cell strainer (Falcon 40μm) and plated at a low density (approximately 400 cells per 10cm dish). After ~14 days, independent colonies were trypsinised with cloning discs (Sigma) and expanded. Knockdown efficiency was tested by qPCR as described below.

***Western blot analysis***

Cells were washed with cold PBS and lysed in 1% SDS (sodium dodecyl sulphate) protein lysis buffer, with addition of proteinase inhibitor and PhosStop solution (Roche) for phosphorylated protein. Protein concentrations were measured using Bio-Rad DC (detergent compatible) protein assay and 40 μg protein in SDS sample buffer and 2-Mercaptoethanol was applied to Mini-PROTEIN TGX Gels 4-20% (Bio-Rad, Hercules, CA, USA), subjected to gel electrophoresis at 115V for 1 hour and 150V for 10 minutes, transferred and blotted to PVDF membranes for 10 minutes at 25V with Trans-Blot Turbo Transfer System (Bio-Rad). Membranes were blocked with Odyssey Blocking Buffer (TBS; LI-COR Bioscience) for 1 hour at room temperature and incubated with the primary antibody (1:500-1:1000 in TBS-T, Supplementary Table S11) overnight at 4°C. After washing the membranes for 3x10 minutes they were incubated with the secondary goat anti-mouse or goat anti-rabbit AB coupled infrared (IR) dye 680 RD or 800 CW (LI-COR, 1:10,000) for 1 hour and, after another 3 washing steps, membranes were imaged using the Odyssey Imaging System (LI-COR).

***RNA extraction and qPCR***

Total RNA was extracted from cells using RNeasy Kits (QIAGEN) with on-column DNase digestion, of which 1 μg was reverse transcribed into cDNA using the SensiFAST cDNA Synthesis Kit (Meridian Bioscience). Transcript abundance was measured by real-time quantitative PCR (qPCR) using the SYBR Green qPCR assay (Applied Biosystems) on the LightCycler 480 (Roche), with each PCR performed in triplicate. Primer sequences are listed in Supplementary Table S12. Gene expression was estimated using the comparative threshold cycle method[21] (delta-delta Ct) against the average Ct value obtained for two control genes (*GAPDH* and *HPRT*).

***Cell viability assay***

Cells were seeded at a density of 1 to 8x$10^3$ per well, depending on growth rates, in 384-well microtiter plates (Corning®) and incubated overnight. Cisplatin (100 µM; Selleck Chemicals) and olaparib (80 µM, Selleck Chemicals) were diluted in 3-fold steps to create a 10-point dose curve; paclitaxel (0.3 µM, Selleck Chemicals) was diluted in 4-fold steps to create a 12-point dose curve. Following 72 hr (cisplatin and paclitaxel) or 120 hr incubation (olaparib), cells were fixed in 2% paraformaldehyde for 10 minutes, washed with PBS and stained with 0.19% Triton X solution containing DAPI (1:1000; SIGMA). Cell dispensing, media changes, and fixing and staining of cells were conducted robotically (BioTek Instruments, Winooski, VT, USA). Drug dispensing was performed with ALH3000 Liquid Handler (PerkinElmer, Waltham, MA, USA). To assess cell viability, the whole area of each well was captured at 10x magnification with CX7-LZR instrument (Thermo Fisher Scientific) and images analysed with CellProfiler v3.0 pipeline. Low quality out-of-focus images (4% of total images) were excluded by manual review before downstream analysis. Non-linear regression drug curves were calculated using GraphPad Prism version 9.3.1 and curve fit compared between *RB1* WT and *RB1* KO clones by an extra sum-of-squares F test.

***Clonogenic survival assay***

Cells (0.8 to 3x$10^3$) were seeded in 6-well plates (Corning®) depending on cell doubling rates. After 12 hours, duplicate wells were treated with cisplatin, paclitaxel or a combination of both drugs at the respective $IC_{50}$ drug concentration, as determined by the 72-hour viability assay.

Cells treated with media alone and with DMF solvent containing media served as controls. After 16 days, cells were rinsed with PBS, fixed and stained with 0.1% crystal violet and methanol for 20 minutes. The whole area of wells was captured in bright-field at 2x magnification using the CX7 (Thermo Fisher Scientific) and the number of clones assessed with the CellProfiler v3.0 software.

***Cell proliferation rates***

Cells were counted using the Countess 3 Automated Cell Counter (Thermo Fisher Scientific) and seeded in 200 µl media in 96-well Corning® plates in triplicate wells and incubated at 37°C. Cells were plated at three different densities (AOCS1 $6x10^3$ to $8x10^3$ cells/well, AOCS7.2 8 to $12x10^3$ cells/well; AOCS16 14 to $18x10^3$ cells/well) according to a previously observed 20% cell confluency per well on day 1, and media changed after 5 days. The whole well area was captured in brightfield every 12 hours for 9 days using real-live cell imaging (Incucyte® Zoom) and cell proliferation rates determined with Incucyte® software. Growth rates were analysed separately in triplicate wells with a starting confluency of between 15% and 25% in three independent experiments.

***Cell cycle profiling***

Cells were seeded in 12-well Corning® plates at between 8 to $12x10^4$ cells/well (AOCS1 $8x10^4$, AOCS7.2 $10x10^4$, AOCS16 $12x10^4$ cells). After 24 hours, each cell line was treated at half the concentration of the respective $IC_{50}$ (determined in the above-described cell viability assay) of either cisplatin (AOCS1: 0.25 µM, AOCS7.2: 0.25 µM; AOCS16: 0.15 µM), paclitaxel (AOCS1: 1.25 nM; AOCS7.2: 50 nM; AOCS16: 0.4 nM) or a combination of both drugs for 24 hours. Cells were rinsed with PBS, trypsinised to form a single-cell suspension and fixed by adding ice-cold 70% ethanol drop-wise. Cells were pelleted and resuspended in a solution containing propidium iodide (0.05 mg/ml) and ribonuclease A (RNase A, Thermo Fisher EN0531, 10 mg/ml). Following 30 to 60 minutes of incubation at room temperature, DNA content was measured using the FACS Canto LSR II flow cytometer. FlowlogicTM software (Inivai) was used to analyse cell cycle distribution in FL3-A channel applying the Watson pragmatic algorithm[22].
